# Understanding the Relational Aspects of Networks of Care for Maternal and Newborn Health: A Revised Operational Framework

**DOI:** 10.1101/2025.01.05.25320026

**Authors:** Olufunke Fasawe, Alisa Jenny, Stefano Bertozzi, Katherine Kalaris, Robyn Churchill, Ndola Prata, Dennis Matheka Mulwa, Marite Vegara, Andy Carmone, Allisyn Moran, Dilys Walker

## Abstract

**Objective:** We investigated the relational aspects of the “Networks of Care” (NOC) approach to better define their role and highlight their importance. These relational aspects distinguish NOC from other health system strengthening approaches that aim to enhance the quality of MNH care. NOCs include functional and relational aspects, as defined in the WHO NOC for MNH framework, and offer a promising strategy for enhancing maternal and newborn health (MNH) by strengthening care coordination at an operational level.

**Methods:** We conducted a cross-disciplinary (healthcare, business, military, aviation, education) scoping review of 90 papers and consulted with key stakeholders to identify and synthesize insights about relational aspects and the associated barriers and facilitators for implementation. We then used the literature review findings and stakeholder feedback to define the relational aspects and revise the Networks of Care Operational framework.

**Results:** The revised framework highlights teamwork with its attributes—trust, mutual respect, shared goals, shared values, and psychological safety and behaviors - leadership, shared decision making, situation monitoring and communication. Organizational enablers underlie a NOC to mitigate barriers and strengthen facilitators to teamwork, which include conflict management, complexity of targets, systemic challenges, social ties, formalization of NOC, and roles like boundary spanners. The study identified relational coordination theory as the basis for refining the NOC operational framework.

**Conclusion:** The resulting framework centers relational coordination including teamwork supported by organizational enablers, in enhancing care coordination between health workers, system administrators, and other users and non-health actors in the network. By targeting relational elements in intervention design and evaluation, the revised Operational NOC Framework for MNH may be valuable in strengthening NOC effectiveness and driving quality improvements leading to improved MNH outcomes, and resilient health systems.

## INTRODUCTION

Despite progress made in reducing maternal and newborn mortality rates globally over the past 30 years, the rates have stagnated since 2015 (1). Access to adequate maternal and newborn health services remains unequal across countries, with sub-Saharan Africa showing the least progress. We are far from achieving the maternal and perinatal SDG goals for 2030, and urgent action is needed (2). While skilled birth attendance is widely available across many countries, most LMICs continue to lack sufficient numbers of health care workers, and maldistribution of healthcare workers is a near universal challenge. This, also with inadequate advanced and referral health services in low-resource settings, mean that improving the coordination of care across various service delivery points and providers, from community-based services to referral centers, is necessary. Efficient coordination of healthcare services involves both the institutional pillars within the healthcare system and the human factors and relational processes that facilitate the delivery of effective, timely, and high-quality care. Unfortunately, insufficient understanding of how MNH services are coordinated through these relational processes across and within multiple service points along the continuum of care has resulted in missed opportunities to improve services, especially in resource-constrained settings.

The Networks of Care (NOCs) approach for maternal and newborn health has become an increasingly important concept in global health. The World Health Organization (WHO) and its collaborators define NOC as “collections of public and/or private health facilities and health workers deliberately interconnected to promote multidisciplinary teamwork and collaborative learning to provide comprehensive, equitable, respectful, person-centered care from home/community to primary through to tertiary levels” (3) NOCs or “purposefully interconnected service delivery touch points” are characterized by coordination across the multiple touchpoints and operational levels of care for mothers and newborns (4). Beginning in 2019, the World Health Organization and others developed a framework to describe the NOC concept as an alternate systems approach to improve maternal and newborn health and strengthen primary healthcare services in low- and middle-income countries where the global burden of maternal and newborn mortality remains very high(3). This multi-partner effort involved WHO, the United States Agency for International Development (USAID), the Bill and Melinda Gates Foundation, the Clinton Health Access Initiative, Jhpiego, and others who have experience with integrated maternal and newborn health programs (5).

While the idea of coordination of services is not new, the NOC approach differentiates itself from conventional service delivery models by emphasizing the “relational” aspects of coordination and recognizing their role in addition to the structural aspects of the healthcare system. These “relational” aspects refer to the often overlooked “soft skills,” that are the human factors or interpersonal and social interactions and processes which drive organizational function. Unfortunately, these relational aspects, generally identified as communication and collaboration, are often assumed to be intact and yet are poorly defined, hindered by ingrained hierarchies, gender, cultural, or other structural barriers. Ultimately, they tend to be neglected but are critical potential levers for improving the effective delivery of healthcare services, including respectful maternity care and patient-focused care. Literature from high-income countries shows that interpersonal, inter-organizational, and intra-organizational relationships are crucial in improving patient care and outcomes (6–8). Several case studies from low- and middle-income countries (LMICs), including Madagascar, Nepal, Nigeria, the Philippines, and Tanzania, studied the elements of NOCs for Maternal and Neonatal Health. These studies highlighted the role of intentional collaboration and communication in establishing and coordinating functional referral systems, promoting community engagement with healthcare facilities, and strengthening service delivery systems. These relational aspects of NOCs were also critical in adapting to challenging or changing conditions toward improving outcomes. The case studies further highlighted the importance of building trust and linkages between healthcare providers at different levels of the health system (9–13).

The original NOC operational framework is theoretical and may be challenging to apply at country and subnational levels for health systems looking to adopt the NOC approach (14). Therefore, there is a need for a more granular understanding of the relational elements of the NOC model framework and its application, particularly for low—and middle-income countries (4,15). By systematically reviewing and understanding relational aspects, we can better define and identify how to apply the NOC approach, measure and act on the relational aspects, and improve MNH service delivery using the NOC approach.

Several systematic reviews have identified key characteristics underlying effective coordination, cooperation, and collaboration. both intra- and inter - organizationally, in healthcare and non-healthcare settings (16–21). Our work builds on these prior reviews, which focused primarily on high-income settings, to synthesize and articulate relational elements that would be applicable in NOCs in Low and Middle Income Countries (LMICs). Thus, this study aims to advance the NOC approach by defining the relational aspects of NOC and their interconnection to other components to inform a revised Operational NOC Framework MNH in LMICs.

## MATERIALS AND METHODS

We used a three-part process: 1) a scoping review of both the published and grey literature, utilizing database searches, reviews of relevant program reports, citations shared by experts, and citations in referenced papers, 2) stakeholder consultations, and 3) development and iteration of a revised framework.

### 1) Scoping review

We conducted a scoping literature review of relational elements and definitions of these elements in relation to organizational networks. Given the nascent literature on networks of care in global health, we also searched other sectors, including business, psychology, sociology, organizational behavior, military, and aviation, in addition to healthcare. We searched for literature exploring the elements or factors associated with improved performance in intra-organizational and inter-organizational collaborations, partnerships, or networks. We searched for all available literature and did not apply limits to the date or geography. We searched – Pubmed, and Scopus. We consulted public health and business experts at the onset and reviewed some preliminary literature referred to us. We used their insights to refine the search strategy and search terms. Following the Joanna Briggs Institute three-step strategy (Aromataris & Munn, 2020), we initially searched the topic ‘*Networks of care’* in Pubmed. Reviewing the common words we identified from titles and abstracts of search results, we generated a list of keywords to refine the search strategy. Applying these keywords, which included inter-organizational, intra-organizational, collaboration, relational coordination, networks, maternal healthcare, teamwork, facilitators, enablers, and barriers, we conducted a more detailed search in the databases. Finally, we reviewed the reference lists and “cited by” articles of the full-text reviewed papers to identify additional papers missing from our search strategy to be included in the study. The search strategy (from Pubmed) was: ("Inter-professional" OR "Interdisciplinary" OR "Inter-organizational collaboration" OR "Intra-organizational collaboration" OR "Inter-organizational relationships" OR Teamwork OR "Effective teamwork" OR "Organizational effectiveness" OR organizational efficiency OR "Relational coordination") AND ( "Network functionality" OR "Network effectiveness" OR "Network relations" OR networks) AND ("health care" OR healthcare OR maternal health services OR "Healthcare organizations" OR organizations OR aviation OR military OR business OR industry) AND (facilitators OR enablers OR barriers).

We excluded systematic reviews from this search but used them to ensure we accessed all relevant literature. Through stakeholder consultations and expert recommendations, we identified additional program reports and grey literature for inclusion.

We conducted an initial title screen of the search outputs, followed by abstract and full article screens. In reviewing the abstracts, we excluded articles that had no direct link with the objectives and scope of the literature review, articles that were study protocols to evaluate organizational relationships or collaboration, and articles that focused on training or education of students for inter and intra-organizational collaboration. We did not include papers that explored relationships between providers and patients. We included all studies published prior to September 2023.

#### Data Extraction

We extracted the data from the retrieved papers into a standardized Excel template. Extracted information including title, authors, doi or website, year of publication, abstract, a summary of the article’s contents, and the relational elements identified. We then synthesized the findings based on commonly occurring themes. We conducted a manual analysis along three dimensions that would inform the revised framework: 1) definitions of relational elements in inter/intra-organizational collaborations or networks, 2) facilitators and barriers of relational elements, and 3) envisioning strategies for improving relational elements.

### 2) Stakeholder consultations

Before conducting the scoping review, we met with colleagues from WHO, USAID, and other stakeholders at the International Maternal Newborn Health Conference in Cape Town in May 2023. Feedback from the stakeholders, a mix of country and global opinion leaders, and policymakers in global health, indicated that conceptually, the concept of a NOC model was clear, but articulating and understanding the relational components and their placement and application in the existing framework was more challenging.

During the summer of 2023, concurrently with the scoping review, our team conducted individual virtual discussions with twenty-one experts specializing in MNH and global health drawn from the WHO-led NOC steering committee and country government officials. They represented a range of organizations and institutions, including the WHO and the World Bank, academic institutions such as Stanford University, bilateral and donor agencies including USAID and the Bill and Melinda Gates Foundation, and various Non-Governmental Organizations (NGOs) and country programs like MOMENTUM, Jhpiego, Institute for Healthcare Improvement (IHI), and CHAI. Thirteen of them represented global organizations, and eight were country-based. We also sought input from two business school professors.

The primary objective of these discussions was to gather diverse perspectives on applying the NOC framework in the LMIC context. The discussions followed the same guide with five questions about their perspectives on the relational processes that make NOCs function and what would be most useful for in-country health system managers in LMICs looking to implement an NOC model.

These interviews with key stakeholders were consultative and did not involve the collection, analysis, or reporting of specific data; thus, the work was exempt from human subject (IRB) review.

### 3) Development of a revised Operational Framework and definitions

Through the scoping review synthesis and the feedback from the MNH experts we consulted, we identified the core relational elements and organizational processes to inform a revised NOC operational framework. The revised framework is conceptualized from an applied perspective that implementers, systems, and policy decision-makers can use to guide interventions and approaches to strengthen NOC. We reviewed existing definitions from a range of sources related to healthcare, and based on expert opinion, we generated definitions that most appropriately describe the relational aspects of the NOC.

## RESULTS

### Scoping Review

The article selection process is summarized in Figure 1. The search yielded 3,569 unique records drawn from online databases and grey literature. We subsequently excluded 3,453 ineligible records through the title and abstract review and fully reviewed 116 records, which included 3 articles found through reference checks. We then assessed these for eligibility and identified a total of 90 eligible articles that were included in the scoping review.

**Fig 1.**
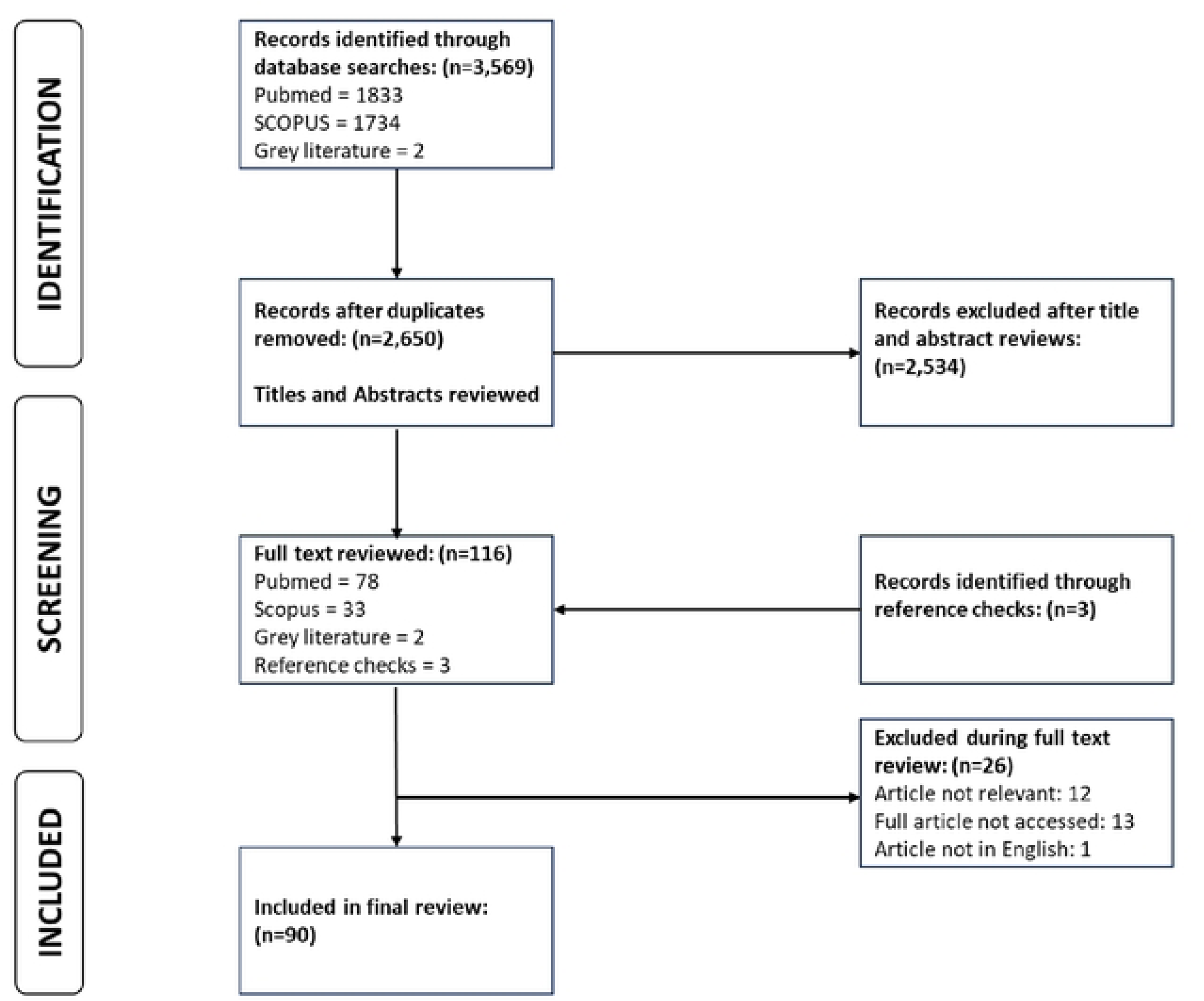
Article Selection and Screening

#### Characteristics of the included articles

Among the reviewed articles, 84% focused on high-income or upper middle-income countries, while only 9% focused on low- and middle-income countries. The low-income and lower middle-income countries represented were Ethiopia, Malawi, Ghana, India, the Philippines and Zambia. The United States and Canada accounted for 50% of all articles from high-income countries.

Eighty-nine of the articles were from the healthcare sector. The majority of the studies (69%) used qualitative methods, followed by quantitative (12%), mixed methods (6%). There has been an increase in the number of articles published on this topic over the past decade. The distribution of the articles by year of publication is shown in Figure 2 and the number of articles by country is shown in Figure 3.

**Fig 2.**
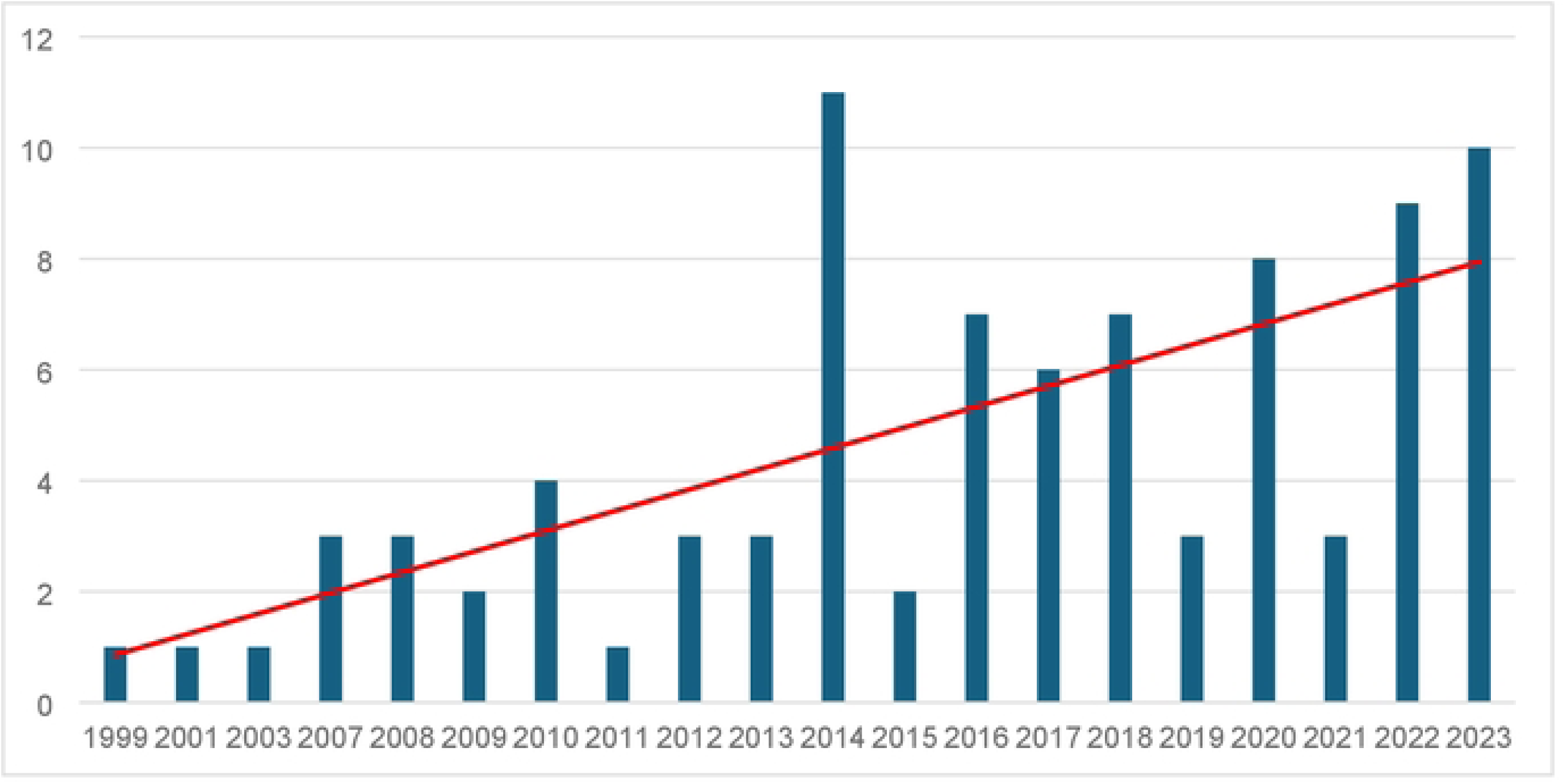
Number of articles reviewed by year of publication (N=90).

**Fig 3.**
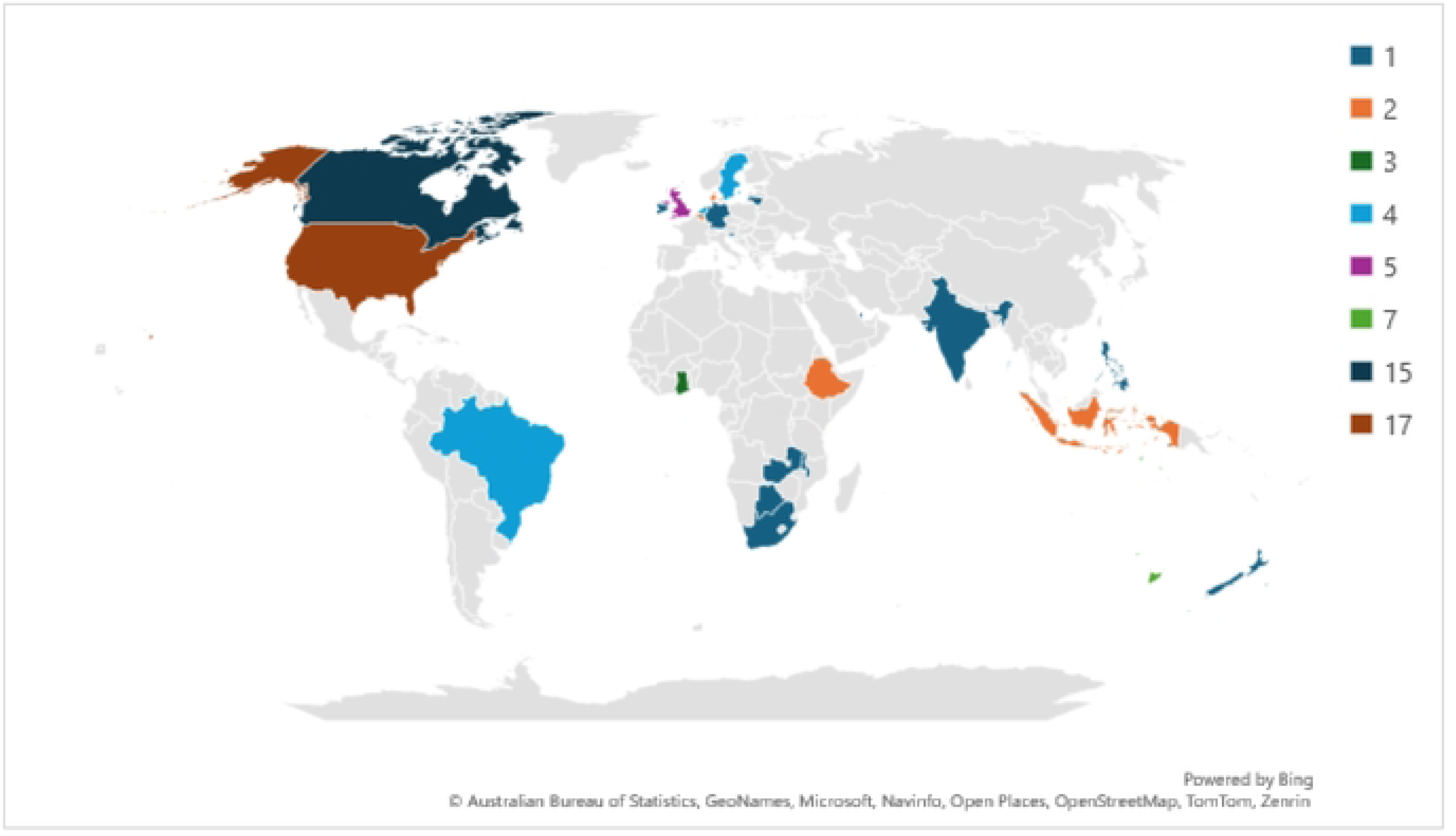
Number of articles reviewed by country.

### Findings of the Scoping Review

#### a. Relational aspects – Attributes and Behaviors

Almost all of the articles referenced trust, mutual respect, shared goals, and shared values as key attributes of relationships or social interactions that make collaboration possible. Trust-building was often cited as a foundational relational element for ensuring effective coordination of collaboratives. Studies emphasized key characteristics of trust, including that partners and stakeholders have a shared knowledge of each other’s roles and expertise and how they contribute jointly to achieving the shared goals of the collaboration (22–24). Linked to trust, some articles highlighted the roles of conflict and conflict management and other key dynamics related to power and hierarchy in influencing the development of trust between collaborating parties (25–29). While critical in forming and maintaining networks, trust is dynamic and non-linear. In addition, to trust, almost all the articles identified the role of mutual respect among collaborators in fostering collaboration and reducing conflicts, leading to greater trust and better communication and teamwork(30).

Most articles reviewed strongly emphasized shared values and a common goal as key components of collaborative engagements that improve coordination. Several of the studies identified the value of aligning toward a shared vision, such as patient-centered care. Patient-centered goals are useful in creating a value proposition for all stakeholders in the network and reduce interpersonal, interprofessional, inter-organizational and intra-organizational tensions that may arise from differences in priorities (24,31–34). A shared mental model, or having a shared goal and vision, enhances trust building, reduces conflict within the collaboration, and, in turn, drives team effectiveness and satisfaction (35). Having a shared mental model focusing on quality healthcare services and patient health outcomes can improve coordination and overall team effectiveness (36).

Another key attribute frequently identified from the review is psychological safety. This builds on interpersonal trust being developed within a team and fosters teamwork in the coordination of care. Articles linked the facilitatory role of leadership to the development of psychological safety, which creates the enabling environment for conflict resolution, deepening of trust, clarity of roles, and feeling supported to take risks and try new things without fear of failure or blame (29,36–39).

A majority of the studies identified behaviors, such as leadership and communication as essential for enhancing coordination and teamwork both within the same units or organizations, or between organizations(40–46). A leader who fosters collaboration, coordination, and shared decision-making was consistently identified as important to effective teamwork (12,47). Leaders’ committing to a collaborative environment and culture was highlighted as important for care quality and coordination(26,31,37,48–50). A leadership structure that supports shared goals, values and vision, fosters teamwork and has mechanisms for managing conflicts and enhancing psychological safety will promote successful collaboration (27,39,50,51). Some of the articles also highlighted the role of leadership, especially institutional leadership, in driving joint planning and coordination across teams and groups of stakeholders in building relationships (22,49).

Most of the articles highlighted effective communication as a key behavior of both inter- and intra-organizational collaboration. Communication between stakeholders and partners should be frequent, open, consistent and foster joint problem-solving, planning, and relationship building (22,28,38,41,52,53). Several articles also identified the importance of creating avenues for communication including regularly scheduled meetings, huddles, or debriefs to facilitate teamwork and cooperation, as these platforms enable the identification and timely resolution of problems thereby reinforcing and promoting the shared commitment to defined outcomes (28,34,36,54–57).

### b. Barriers and facilitators of relational elements

Many articles identified barriers and facilitators that influence the relational elements necessary for optimal collaboration and coordination. These barriers and facilitators are broad and diverse ranging from interpersonal relationships and conflicts; to a poor understanding of, or overly complex goals; to systemic or structural barriers that hinder collaboration. These are presented in greater detail in Table 1.

**Table 1.** Barriers and facilitators to relational elements in Networks of Care.

| Barrier | Description | Facilitator | Description |
| --- | --- | --- | --- |
| Unresolved conflicts | Can weaken interpersonal relationships and lead to erosion of trust. Conflict in interdisciplinary healthcare teams, though often inevitable, can democratize relationships and spark creativity. It provides an opportunity for diverse voices to be heard, promoting shared power in decision making and collective autonomy (25,56,58–60). Team meetings were emphasized as a crucial arena for addressing conflicts. | Interpersonal relationships and social ties | Social ties and interpersonal relationships play an important role in collaboration. A lack of familiarity and pre-existing personal relationships affected the development of trust and respect in the collaboration between clinical and community-based social service organizations (61). |
| Lack of a mutual understanding of the purpose of the collaboration | Without a common and clearly defined shared purpose (shared goals), there is lack of cohesion threatening shared ownership for the desired results of the collaboration (47,62) | Formalized partnerships | Establishing formal agreements such as MOUs, terms of reference, and other governance documents serve as a foundational structure that bolsters and reinforces trust between partnering entities. This scaffolding mechanism aids in achieving common goals, objectives, and mutually beneficial outcomes(12,23,28,42,63–66). |
| Overly complex goals and targets | At an organizational level, setting complex goals and targets that are unachievable could dampen motivation among stakeholders | Boundary spanning roles | The role of “boundary spanners” or staff who work across different boundaries (organizational, professional, geographical) was highlighted. These boundary spanners were noted as necessary for maintaining commitments towards the shared goals for the collaborative efforts across the different groups in the network (25,26,31,67–69). |
| Systemic barriers | Includes staffing shortages or high turnover which contribute to staff overload and inadequate time to participate in activities that promote coordination; lack of institutional leadership support, inadequate funding, and lack of relevant supporting infrastructure e.g. communications are also important (26,41,64,70–73) | Geographic or physical proximity | Co-location of stakeholders or team members can foster or accelerate the process of developing team shared mental models and commitments to defined shared values and goals for the collaboration (45,61,69,74–76).<br>Co-location of team members also contributes to frequent, timely and accurate communication and promotes joint problem solving and shared decision making (21,26,59,70,74,75,77). |

### Networks of Care Publication – Manuscript draft

The nine articles from low-income and lower-middle-income countries were more likely to identify systemic barriers to collaborative relationships and care coordination. The “Networks of Practice” approach implemented in Ghana highlighted key systemic and structural issues, such as weak health system governance, lack of political will, and poor healthcare financing, which may impact the development and sustainability of the relational elements. Although not extensively identified, power dynamics and hierarchies (especially between physicians and other professionals) (26,57,60,68,75) and less identified, gender (78) were additional barriers of note.

#### c. Strategies to promotteamwork

Some of the studies assessed or described strategies that have been implemented to enhance teamwork and coordination. The majority of these strategies were organizational strategies. Several of the organizational strategies were recurrent across studies and can be grouped into three main categories:1) Role clarity and standardized processes, 2) Performance management systems, and 3) Coordination processes.

##### Role clarity and standardized policies

Some of the studies described formalized processes for information sharing, planning, and problem-solving around a shared goal. Some studies identified the need to establish clear frameworks to ensure stakeholders are aligned on the shared goal, purpose, and expected outcomes for the collaboration(12,24,29,44,47,79–81). These studies also identified the need to define roles and responsibilities. This includes clear job descriptions and adoption of standardized tools, protocols, checklists and standard operating procedures (SOPs) which can help reduce the variation in service delivery and improve coordination between stakeholders.

##### Performance Management

Studies described the need for a management framework that defines the mechanisms for collaboration and expectations for all stakeholders, accountability arrangements, and measures to assess whether the collaborative effort is yielding the desired results. Several studies identified decision-making and accountability arrangements as key factors that contribute to the success of collaborative efforts (12,31,32,79,82).

Some studies identified the importance of applying organizational performance tools such as Lean management or Plan-Do-Study-Act (PDSA) to conduct gap analyses and identify solutions to improve care coordination(77). Lean management has been found to positively impact improving teamwork, communication, and coordination among health workers(83,84). Kerrissey et al. expanded on the need for instituting processes for measuring performance, addressing conflict, correcting mistakes in a timely fashion, and standardizing how these performance audits are done (61).

Other strategies identified include setting measurable targets, measuring them, and rewarding performance (18) as well as focusing on the performance of teams instead of individuals (85). Mechanisms to maintain team stability which helps to strengthen social ties and interpersonal relationships were cited as crucial to enhancing the performance of collaborative engagements. This applies to both intra-organizational teams and inter-organizational teams which make up the NOC. Some articles highlighted the importance of having accountability systems that enable the collaborating group to keep track of progress towards set goals. These processes include conducting audits (74), performance measurement of the collaborative and not the individuals or mutual performance monitoring (18,82,85–87).

##### Coordination

Almost all the studies highlighted three main strategies to enhance coordination. These include: 1) Creating opportunities for physical connections and co-location. These could be through meetings, huddles, and other engagements (44,59,61,64,88,89); 2) Providing information and communication technologies to enhance communication and frequency of communication(90–92); and 3) Identifying and assigning a coordination agent. The individual ensures that processes in place are working and identifies issues that may emerge or need addressing(81,93).

Despite the exclusion of articles specifically describing training approaches, we identified training as a critical component to strengthen relationships among team members, improve collaborative leadership, and enhance coordination. Strategies identified for strengthening teamwork at the relational level include team training, leadership training, and formalized communication tools and training. Some studies identified the role of intentionally strengthening leadership capacity to foster teamwork and build trust and transparency(61). This includes building leadership capacity in teamwork, collaboration, shared decision-making, and network oversight. Studies found that leadership involvement in identifying problems and shared decision-making to identify and implement solutions can strengthen collaborative relationships in healthcare service delivery (82). Mentoring by teamwork specialists provides an effective way to build capacity in teamwork alongside professional skills and competencies.

The need to train staff in both task expertise and teamwork was often highlighted as essential in most articles. According to the research conducted by Wise et al., the effectiveness of a team is not solely dependent on the individuals’ skills and competence to carry out their designated task but also on their ability to coordinate with each other (18,43,76,85). The Team Strategies and Tools to Enhance Performance and Patient Safety (TeamSTEPPS) training is one patient safety training program designed to strengthen teamwork. TeamSTEPPS aims to enhance communication and teamwork in healthcare settings by providing healthcare professionals with the necessary tools and strategies to work collaboratively towards a common goal (94,95).

### Revised operational framework and definitions

In the review, we came across popular frameworks that have been used to assess integration, collaboration, and coordination including the network theory (96), and the competing values frameworks(97). However, the Relational Coordination (RC) theory (77,98) stood out among the theories we identified and appears to be the most suited as a basis for the relational elements of NOCs for LMICs.

Effective communication and coordination are crucial for achieving desired outcomes in work that is highly interdependent and time-sensitive, according to the RC theory developed by Joy Hoffer Gittell in the mid-1990s based on an in-depth field study of flight departures in the airline industry(77)( Fig 4). It expanded on previous work done on coordination. The theory of relational coordination proposes that “relationships with shared goals, shared knowledge, and mutual respect are critical to support frequent, timely, accurate, problem-solving communication, and vice versa, enabling stakeholders to effectively coordinate their work across boundaries” (77). Shared goals, mutual respect and enhanced communication were found to be foundational among effective Networks of Care in the 2020 scoping review and case studies,(4) hence the Relational Coordination Theory is in agreement with previous literature about NOC.

**Figure 4.**
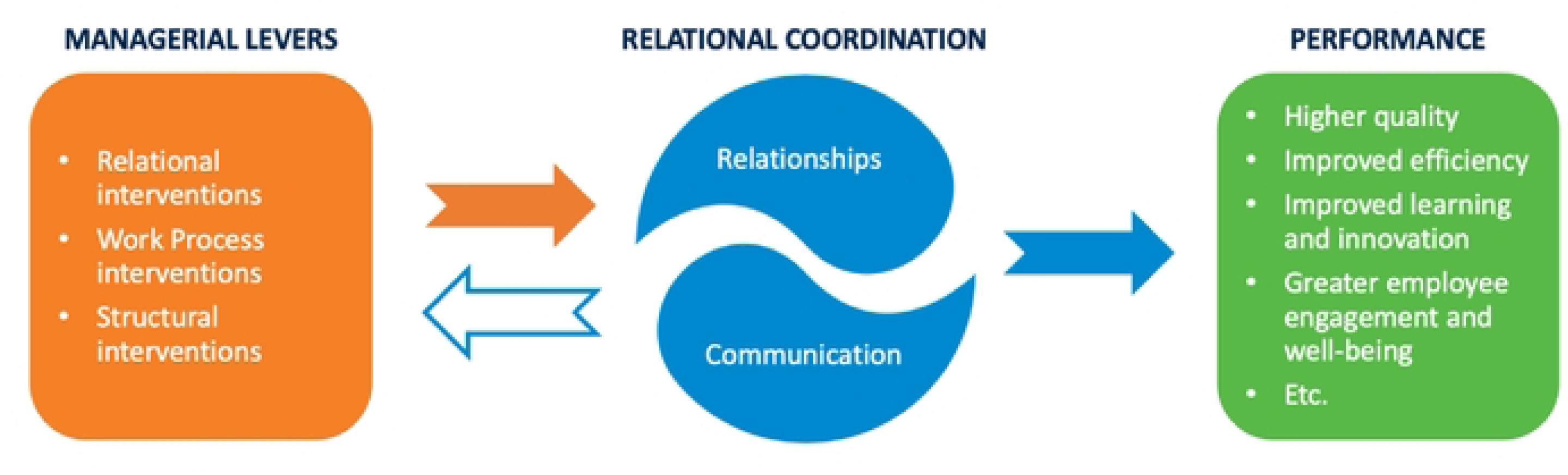
Interventions to strengthen Relational Coordination (Source: https://cisr.mit.edu/publication/2022_0501_RelationalEcosystems_SebastianGittell#fn_4)

When RC is applied to healthcare, studies suggest it can lead to improvements in the quality of care, increased provider job satisfaction, patient-reported care continuity, and adaptive learning (6,99–102).

Including the synthesis of the literature review and the feedback from the MNH experts at the global and country levels, we adapted the relational coordination theory to develop a revised Operational NOC Framework for MNH (Fig 5). This revised framework takes an applied perspective, aiming to serve as a guide that implementers, systems, and policy decision-makers can use to establish and strengthen NOCs. The revised framework, building on the original framework, has two main aspects for a functional NOC model – 1) Structure – made up of the WHO health systems building blocks) and 2) Relational Coordination, subdivided into teamwork and organizational enablers.

**Figure 5.**
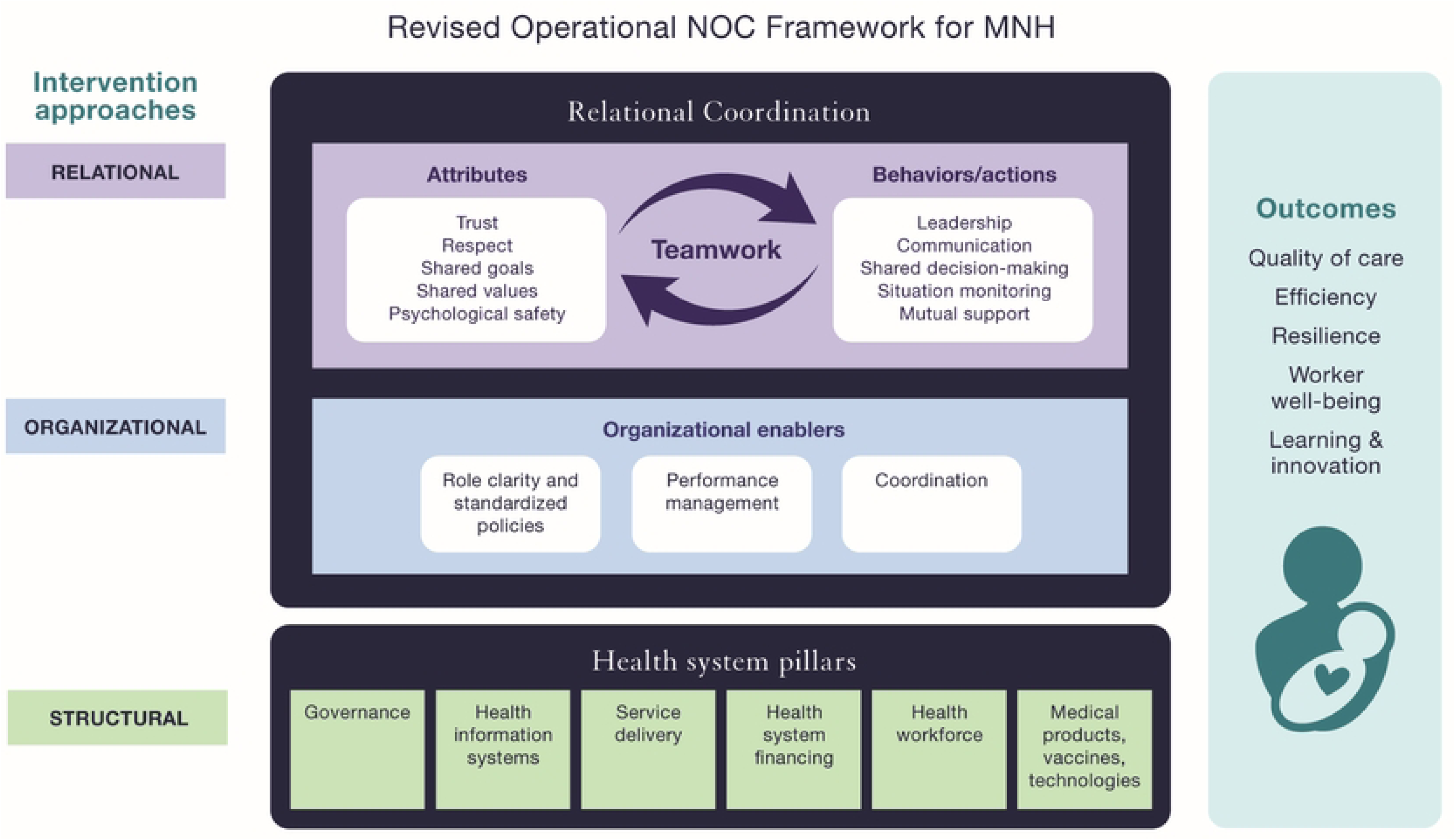
Revised Operational NOC Framework for MNH

### The Revised Operational NOC Framework for MNH

Synthesizing the concepts of teamwork, organizational enablers, and the health system pillars that make NOCs operational and functional, we developed a revised Operational Framework for NOCs emphasizing the relational aspects and how they layer on the structural aspects. Figure 5 illustrates the revised framework we developed. In this framework, Relational Coordination for NOC includes teamwork and organizational enablers within dynamic and iterative processes that encourage adaptive learning and lead to improved performance or outcomes.

After developing the revised framework with some iterations based on stakeholder feedback, we felt it was necessary to modify the NOC definition to reflect better the dynamic processes and mechanisms by which an effective NOC can drive improved outcomes. This includes acknowledging the role of formal and informal relationships and interdisciplinary teams and is proposed as follows:

*Networks of care for maternal and newborn health exist on a spectrum from informal or formal collaborations between and within public and private health facilities interconnected to form interdisciplinary teams with structural, organizational, relational and adaptive learning to deliver high quality equitable, respectful person-centered care from household to primary through to tertiary levels*.

### Defining the Teamwork attributes and behaviors

Our review and consequent operational framework depict teamwork as a dynamic state in which a set of attributes and behaviors are mutually reinforcing. We reviewed existing definitions of the identified attributes and behaviors that comprise teamwork and, through expert opinion and review, generated definitions that most appropriately describe the attributes and behaviors of teamwork in the context of NOC. (Table 2).

**Table 2.** Teamwork attributes and behaviors – Definitions (synthesis from multiple online and published sources relevant to healthcare)

|  | Definition |
| --- | --- |
| **Attributes** |  |
| Trust | The predictive understanding of another's behavior. Shared willingness of team members to be vulnerable to the actions of other members. Required for psychological safety. |
| Respect | The belief that all persons have unconditional intrinsic value as human beings (dignity) and team members and act in light of that belief. |
| Shared Values | Beliefs that are jointly held among members of a team which guide individual and interdependent team behavior and guide decision making. |
| Shared goals | Objectives or outcomes that are collectively determined, agreed upon by a group and which each individual in the group works towards achieving |
| Psychological safety | The willingness of team members to take interpersonal risks, for example by admitting an error, asking a question, or seeking help. |
| **Behaviors** |  |
| Leadership | The practice of influencing team members through effective communication to foster psychological safety to achieve a shared purpose under conditions of uncertainty. |
| Communication | The exchange of information between two or more team members in the prescribed manner and using proper terminology. Often the purpose of communication is to clarify or acknowledge the receipt of information. |
| Mutual support | Team members (1) assisting one another; (2) providing and receiving feedback; and (3) exerting assertive and advocacy behaviors when patient safety is threatened. The essence of teamwork. |
| Situation monitoring | Knowing what is happening in the changing environment and using new knowledge to adapt to the situation and to support team members. |
| Shared decision making | A process that draws on the combined knowledge of all team members to make smarter, more effective decisions. |

**Table 3.**
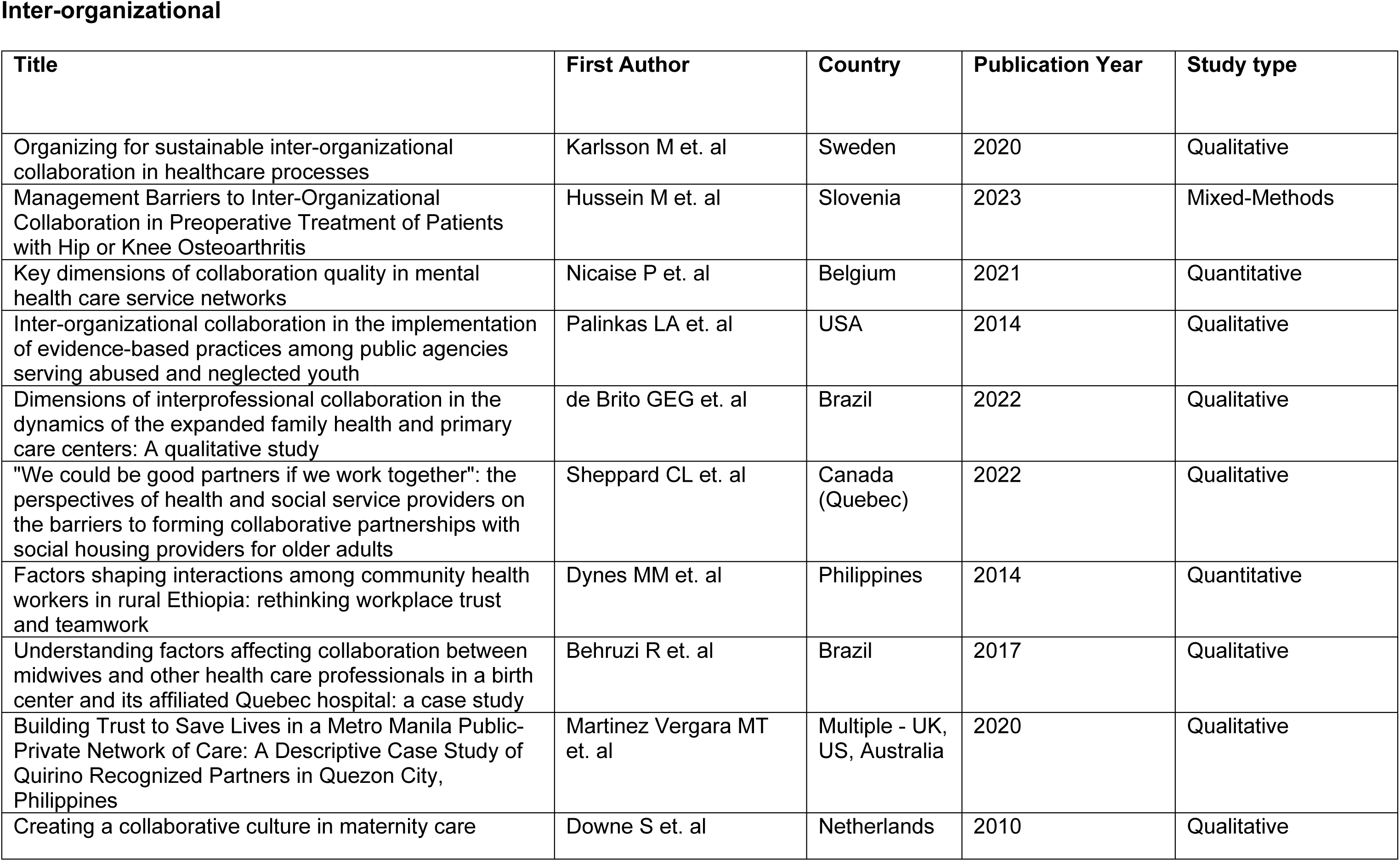
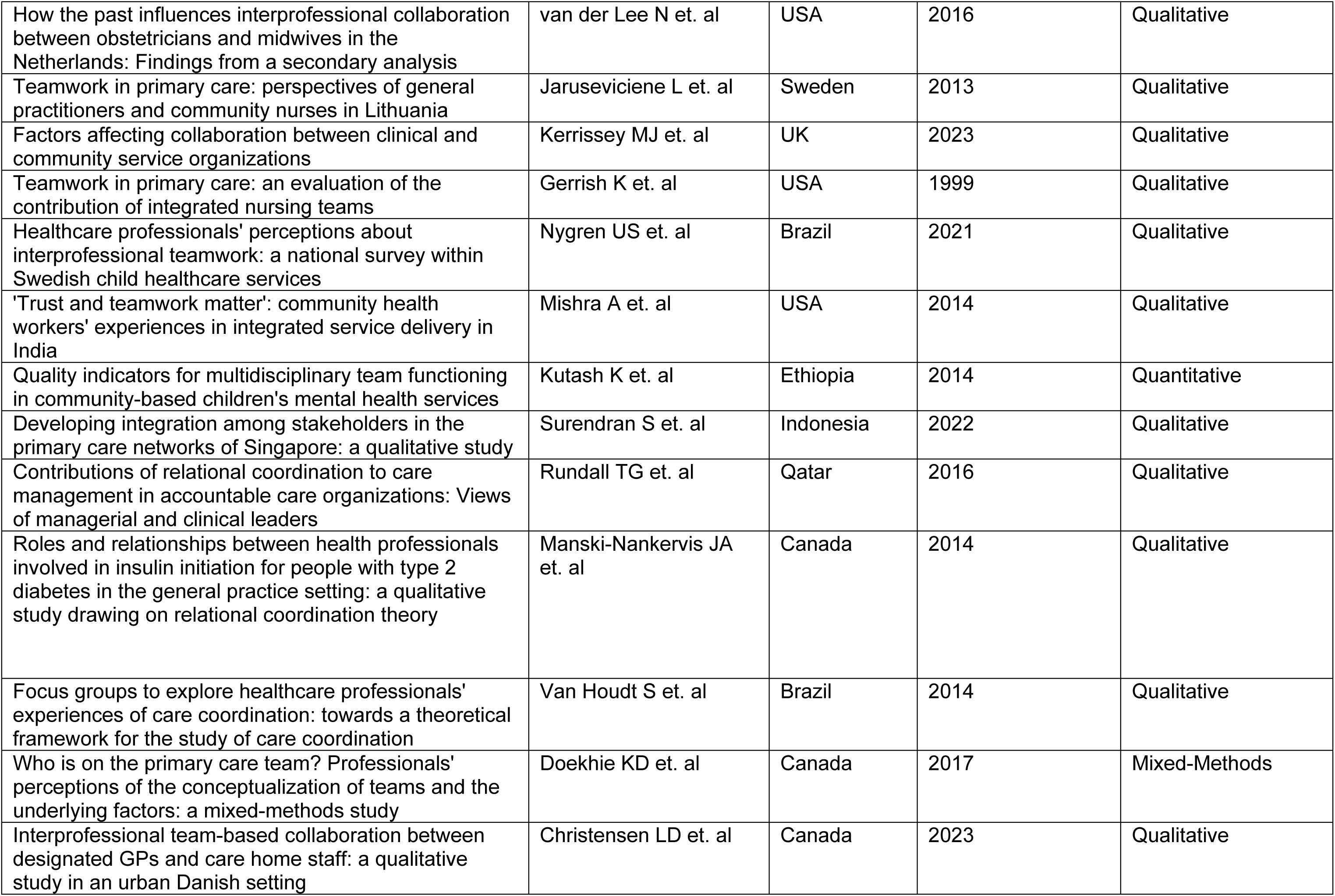
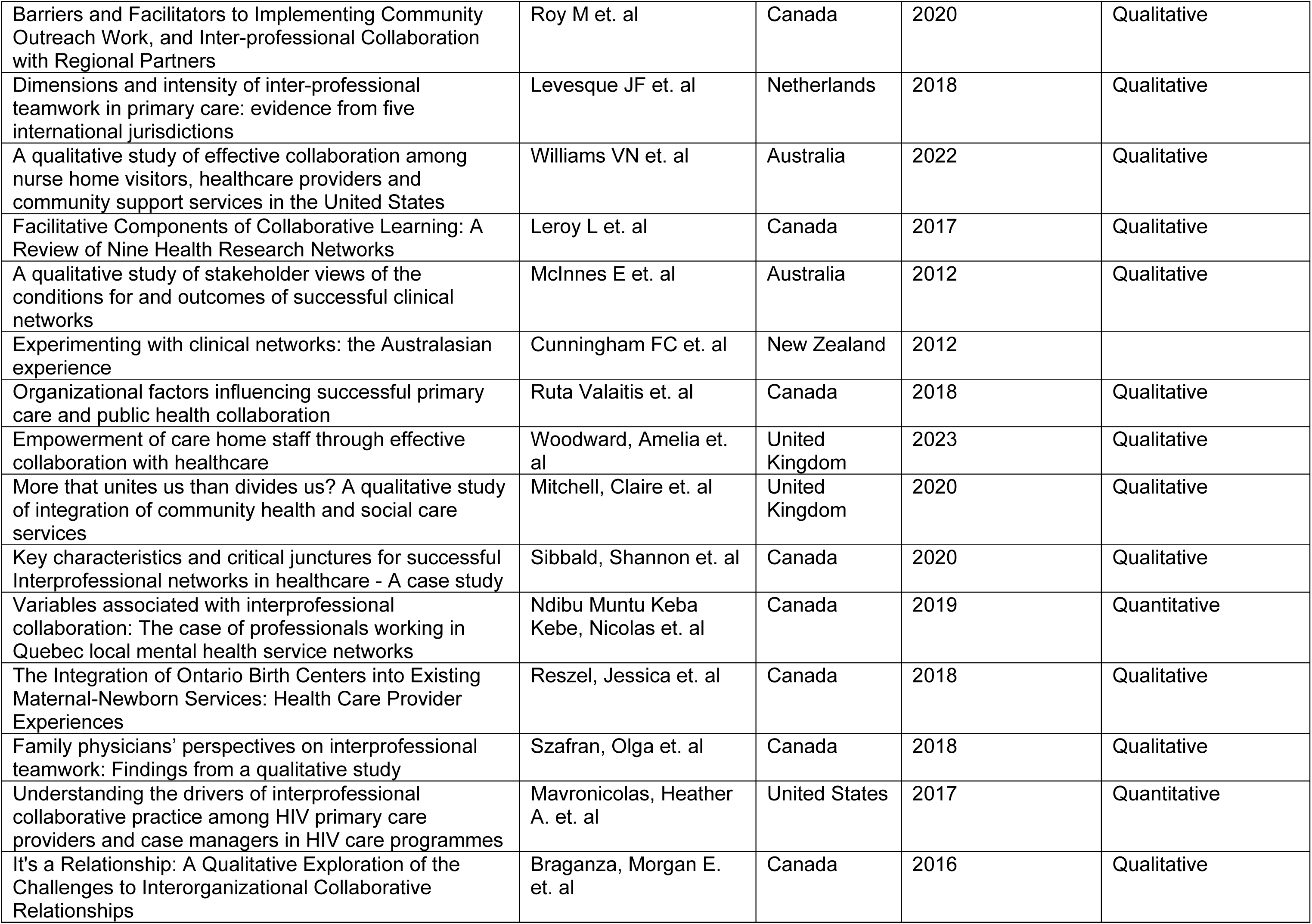
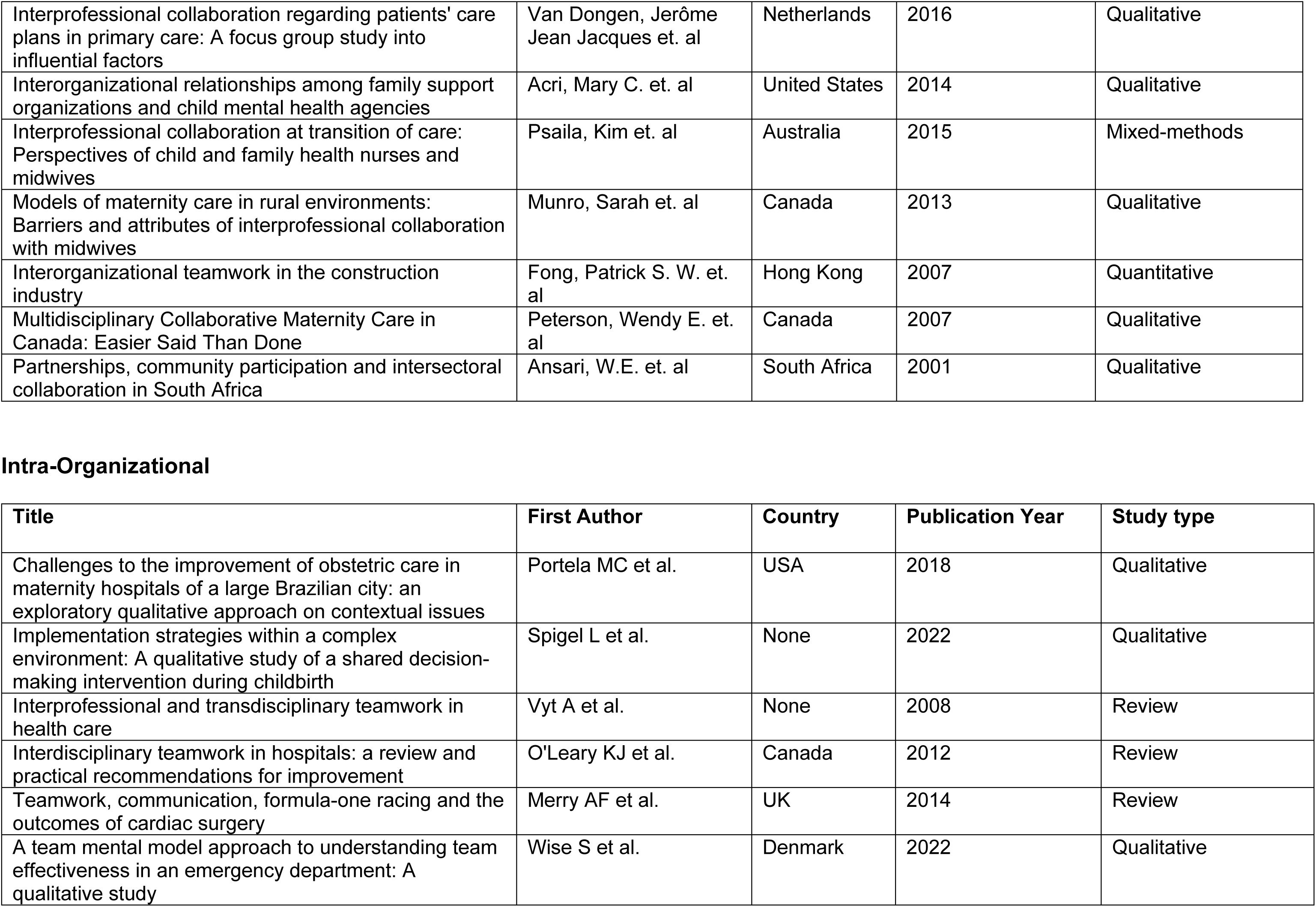
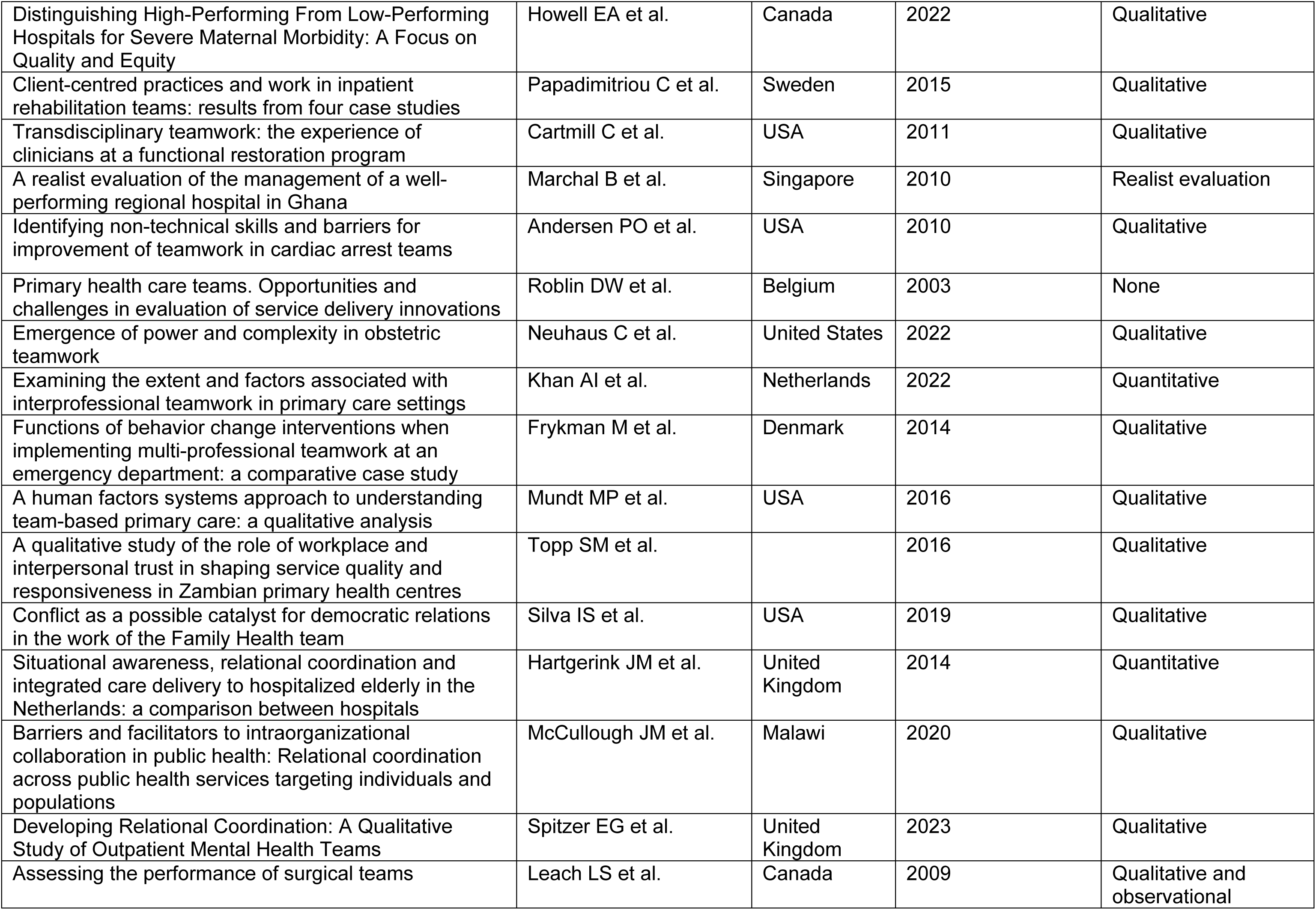
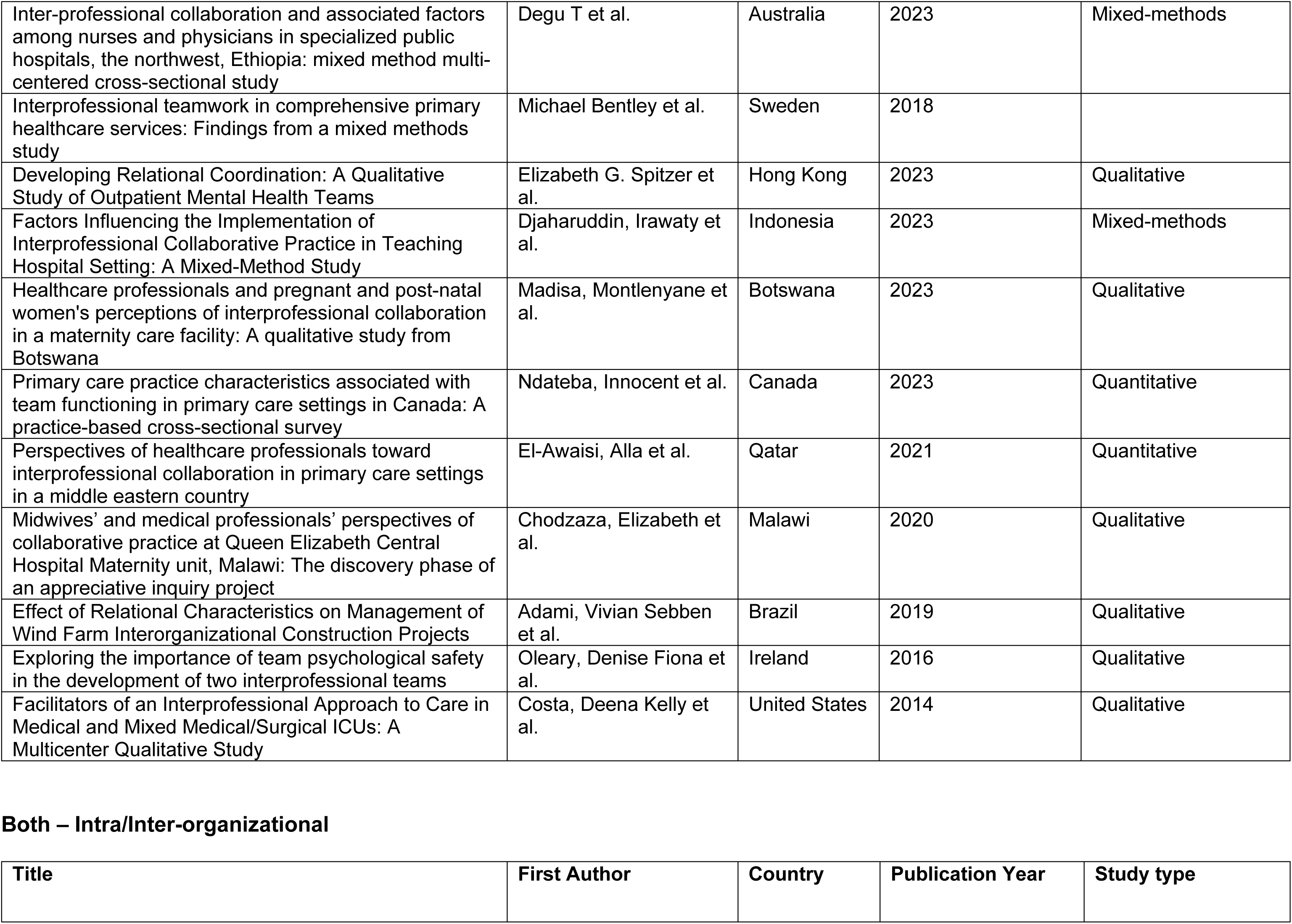
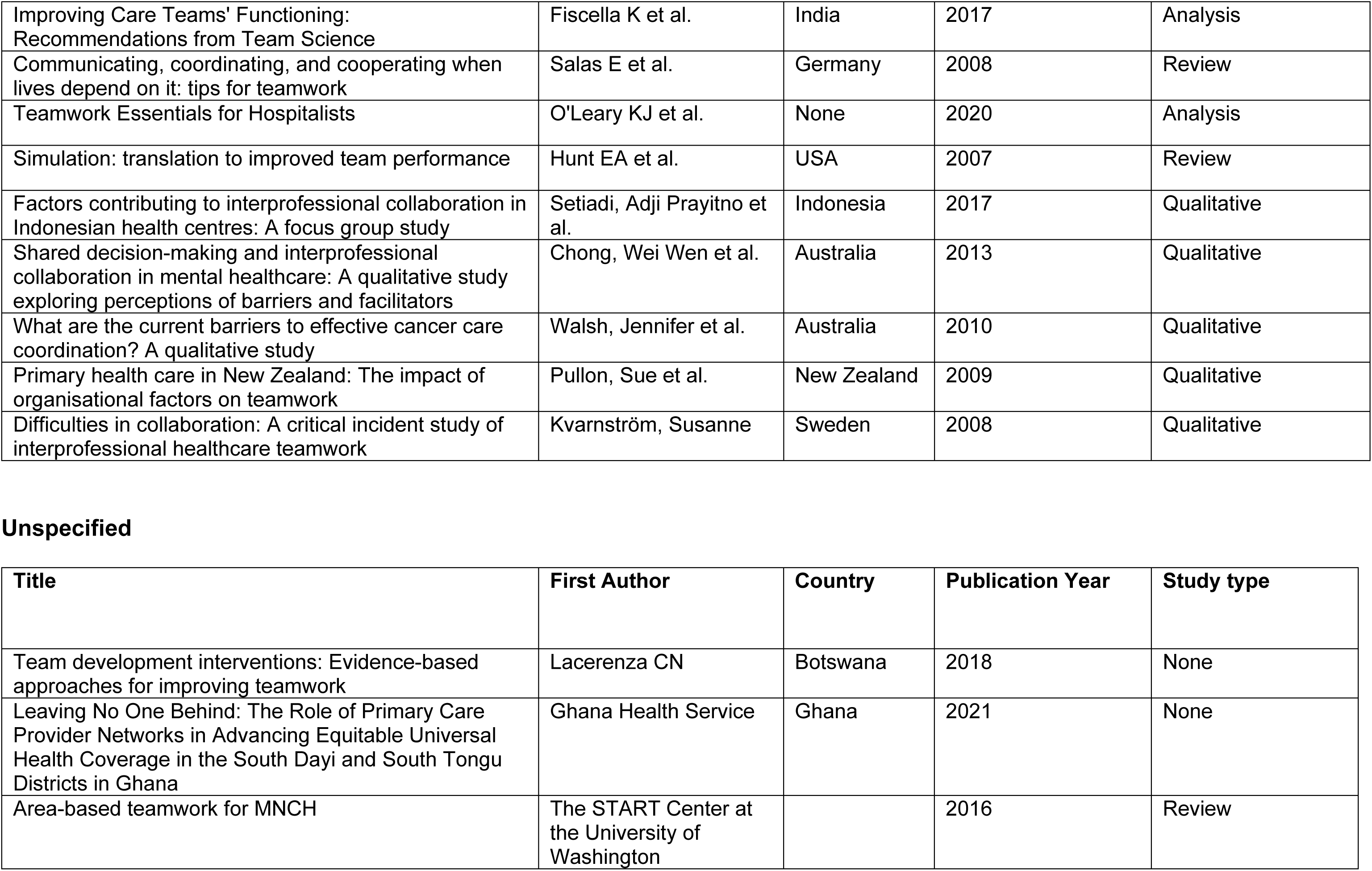
Papers included in the final scoping review.

**Inter-organizational**
| Title | First Author | Country | Publication Year | Study type |
| --- | --- | --- | --- | --- |
| Organizing for sustainable inter-organizational collaboration in healthcare processes | Karlsson M et. al | Sweden | 2020 | Qualitative |
| Management Barriers to Inter-Organizational Collaboration in Preoperative Treatment of Patients with Hip or Knee Osteoarthritis | Hussein M et. al | Slovenia | 2023 | Mixed-Methods |
| Key dimensions of collaboration quality in mental health care service networks | Nicaise P et. al | Belgium | 2021 | Quantitative |
| Inter-organizational collaboration in the implementation of evidence-based practices among public agencies serving abused and neglected youth | Palinkas LA et. al | USA | 2014 | Qualitative |
| Dimensions of interprofessional collaboration in the dynamics of the expanded family health and primary care centers: A qualitative study | de Brito GEG et. al | Brazil | 2022 | Qualitative |
| "We could be good partners if we work together": the perspectives of health and social service providers on the barriers to forming collaborative partnerships with social housing providers for older adults | Sheppard CL et. al | Canada (Quebec) | 2022 | Qualitative |
| Factors shaping interactions among community health workers in rural Ethiopia: rethinking workplace trust and teamwork | Dynes MM et. al | Philippines | 2014 | Quantitative |
| Understanding factors affecting collaboration between midwives and other health care professionals in a birth center and its affiliated Quebec hospital: a case study | Behruzi R et. al | Brazil | 2017 | Qualitative |
| Building Trust to Save Lives in a Metro Manila Public-Private Network of Care: A Descriptive Case Study of Quirino Recognized Partners in Quezon City, Philippines | Martinez Vergara MT et. al | Multiple - UK, US, Australia | 2020 | Qualitative |
| Creating a collaborative culture in maternity care | Downe S et. al | Netherlands | 2010 | Qualitative |
| Networks of Care Publication – Manuscript draft |  |  |  |  |
| How the past influences interprofessional collaboration between obstetricians and midwives in the Netherlands: Findings from a secondary analysis | van der Lee N et. al | USA | 2016 | Qualitative |
| Teamwork in primary care: perspectives of general practitioners and community nurses in Lithuania | Jaruseviciene L et. al | Sweden | 2013 | Qualitative |
| Factors affecting collaboration between clinical and community service organizations | Kerrissey MJ et. al | UK | 2023 | Qualitative |
| Teamwork in primary care: an evaluation of the contribution of integrated nursing teams | Gerrish K et. al | USA | 1999 | Qualitative |
| Healthcare professionals' perceptions about interprofessional teamwork: a national survey within Swedish child healthcare services | Nygren US et. al | Brazil | 2021 | Qualitative |
| 'Trust and teamwork matter': community health workers' experiences in integrated service delivery in India | Mishra A et. al | USA | 2014 | Qualitative |
| Quality indicators for multidisciplinary team functioning in community-based children's mental health services | Kutash K et. al | Ethiopia | 2014 | Quantitative |
| Developing integration among stakeholders in the primary care networks of Singapore: a qualitative study | Surendran S et. al | Indonesia | 2022 | Qualitative |
| Contributions of relational coordination to care management in accountable care organizations: Views of managerial and clinical leaders | Rundall TG et. al | Qatar | 2016 | Qualitative |
| Roles and relationships between health professionals involved in insulin initiation for people with type 2 diabetes in the general practice setting: a qualitative study drawing on relational coordination theory | Manski-Nankervis JA et. al | Canada | 2014 | Qualitative |
| Focus groups to explore healthcare professionals' experiences of care coordination: towards a theoretical framework for the study of care coordination | Van Houdt S et. al | Brazil | 2014 | Qualitative |
| Who is on the primary care team? Professionals' perceptions of the conceptualization of teams and the underlying factors: a mixed-methods study | Doekhie KD et. al | Canada | 2017 | Mixed-Methods |
| Interprofessional team-based collaboration between designated GPs and care home staff: a qualitative study in an urban Danish setting | Christensen LD et. al | Canada | 2023 | Qualitative |
| Networks of Care Publication – Manuscript draft |  |  |  |  |
| Barriers and Facilitators to Implementing Community Outreach Work, and Inter-professional Collaboration with Regional Partners | Roy M et. al | Canada | 2020 | Qualitative |
| Dimensions and intensity of inter-professional teamwork in primary care: evidence from five international jurisdictions | Levesque JF et. al | Netherlands | 2018 | Qualitative |
| A qualitative study of effective collaboration among nurse home visitors, healthcare providers and community support services in the United States | Williams VN et. al | Australia | 2022 | Qualitative |
| Facilitative Components of Collaborative Learning: A Review of Nine Health Research Networks | Leroy L et. al | Canada | 2017 | Qualitative |
| A qualitative study of stakeholder views of the conditions for and outcomes of successful clinical networks | McInnes E et. al | Australia | 2012 | Qualitative |
| Experimenting with clinical networks: the Australasian experience | Cunningham FC et. al | New Zealand | 2012 |  |
| Organizational factors influencing successful primary care and public health collaboration | Ruta Valaitis et. al | Canada | 2018 | Qualitative |
| Empowerment of care home staff through effective collaboration with healthcare | Woodward, Amelia et. al | United Kingdom | 2023 | Qualitative |
| More that unites us than divides us? A qualitative study of integration of community health and social care services | Mitchell, Claire et. al | United Kingdom | 2020 | Qualitative |
| Key characteristics and critical junctures for successful Interprofessional networks in healthcare - A case study | Sibbald, Shannon et. al | Canada | 2020 | Qualitative |
| Variables associated with interprofessional collaboration: The case of professionals working in Quebec local mental health service networks | Ndibu Muntu Keba Kebe, Nicolas et. al | Canada | 2019 | Quantitative |
| The Integration of Ontario Birth Centers into Existing Maternal-Newborn Services: Health Care Provider Experiences | Reszel, Jessica et. al | Canada | 2018 | Qualitative |
| Family physicians' perspectives on interprofessional teamwork: Findings from a qualitative study | Szafran, Olga et. al | Canada | 2018 | Qualitative |
| Understanding the drivers of interprofessional collaborative practice among HIV primary care providers and case managers in HIV care programmes | Mavronicolas, Heather A. et. al | United States | 2017 | Quantitative |
| It's a Relationship: A Qualitative Exploration of the Challenges to Interorganizational Collaborative Relationships | Braganza, Morgan E. et. al | Canada | 2016 | Qualitative |
| Networks of Care Publication – Manuscript draft |  |  |  |  |
| Interprofessional collaboration regarding patients' care plans in primary care: A focus group study into influential factors | Van Dongen, Jérôme Jean Jacques et. al | Netherlands | 2016 | Qualitative |
| Interorganizational relationships among family support organizations and child mental health agencies | Acri, Mary C. et. al | United States | 2014 | Qualitative |
| Interprofessional collaboration at transition of care: Perspectives of child and family health nurses and midwives | Psaila, Kim et. al | Australia | 2015 | Mixed-methods |
| Models of maternity care in rural environments: Barriers and attributes of interprofessional collaboration with midwives | Munro, Sarah et. al | Canada | 2013 | Qualitative |
| Interorganizational teamwork in the construction industry | Fong, Patrick S. W. et. al | Hong Kong | 2007 | Quantitative |
| Multidisciplinary Collaborative Maternity Care in Canada: Easier Said Than Done | Peterson, Wendy E. et. al | Canada | 2007 | Qualitative |
| Partnerships, community participation and intersectoral collaboration in South Africa | Ansari, W.E. et. al | South Africa | 2001 | Qualitative |

Intra-Organizational
| Title | First Author | Country | Publication Year | Study type |
| --- | --- | --- | --- | --- |
| Challenges to the improvement of obstetric care in maternity hospitals of a large Brazilian city: an exploratory qualitative approach on contextual issues | Portela MC et al. | USA | 2018 | Qualitative |
| Implementation strategies within a complex environment: A qualitative study of a shared decision-making intervention during childbirth | Spigel L et al. | None | 2022 | Qualitative |
| Interprofessional and transdisciplinary teamwork in health care | Vyt A et al. | None | 2008 | Review |
| Interdisciplinary teamwork in hospitals: a review and practical recommendations for improvement | O'Leary KJ et al. | Canada | 2012 | Review |
| Teamwork, communication, formula-one racing and the outcomes of cardiac surgery | Merry AF et al. | UK | 2014 | Review |
| A team mental model approach to understanding team effectiveness in an emergency department: A qualitative study | Wise S et al. | Denmark | 2022 | Qualitative |
| Networks of Care Publication – Manuscript draft |  |  |  |  |
| Distinguishing High-Performing From Low-Performing Hospitals for Severe Maternal Morbidity: A Focus on Quality and Equity | Howell EA et al. | Canada | 2022 | Qualitative |
| Client-centred practices and work in inpatient rehabilitation teams: results from four case studies | Papadimitriou C et al. | Sweden | 2015 | Qualitative |
| Transdisciplinary teamwork: the experience of clinicians at a functional restoration program | Cartmill C et al. | USA | 2011 | Qualitative |
| A realist evaluation of the management of a well-performing regional hospital in Ghana | Marchal B et al. | Singapore | 2010 | Realist evaluation |
| Identifying non-technical skills and barriers for improvement of teamwork in cardiac arrest teams | Andersen PO et al. | USA | 2010 | Qualitative |
| Primary health care teams. Opportunities and challenges in evaluation of service delivery innovations | Roblin DW et al. | Belgium | 2003 | None |
| Emergence of power and complexity in obstetric teamwork | Neuhaus C et al. | United States | 2022 | Qualitative |
| Examining the extent and factors associated with interprofessional teamwork in primary care settings | Khan AI et al. | Netherlands | 2022 | Quantitative |
| Functions of behavior change interventions when implementing multi-professional teamwork at an emergency department: a comparative case study | Frykman M et al. | Denmark | 2014 | Qualitative |
| A human factors systems approach to understanding team-based primary care: a qualitative analysis | Mundt MP et al. | USA | 2016 | Qualitative |
| A qualitative study of the role of workplace and interpersonal trust in shaping service quality and responsiveness in Zambian primary health centres | Topp SM et al. |  | 2016 | Qualitative |
| Conflict as a possible catalyst for democratic relations in the work of the Family Health team | Silva IS et al. | USA | 2019 | Qualitative |
| Situational awareness, relational coordination and integrated care delivery to hospitalized elderly in the Netherlands: a comparison between hospitals | Hartgerink JM et al. | United Kingdom | 2014 | Quantitative |
| Barriers and facilitators to intraorganizational collaboration in public health: Relational coordination across public health services targeting individuals and populations | McCullough JM et al. | Malawi | 2020 | Qualitative |
| Developing Relational Coordination: A Qualitative Study of Outpatient Mental Health Teams | Spitzer EG et al. | United Kingdom | 2023 | Qualitative |
| Assessing the performance of surgical teams | Leach LS et al. | Canada | 2009 | Qualitative and observational |
| Networks of Care Publication – Manuscript draft |  |  |  |  |
| Inter-professional collaboration and associated factors among nurses and physicians in specialized public hospitals, the northwest, Ethiopia: mixed method multi-centered cross-sectional study | Degu T et al. | Australia | 2023 | Mixed-methods |
| Interprofessional teamwork in comprehensive primary healthcare services: Findings from a mixed methods study | Michael Bentley et al. | Sweden | 2018 |  |
| Developing Relational Coordination: A Qualitative Study of Outpatient Mental Health Teams | Elizabeth G. Spitzer et al. | Hong Kong | 2023 | Qualitative |
| Factors Influencing the Implementation of Interprofessional Collaborative Practice in Teaching Hospital Setting: A Mixed-Method Study | Djaharuddin, Irawaty et al. | Indonesia | 2023 | Mixed-methods |
| Healthcare professionals and pregnant and post-natal women's perceptions of interprofessional collaboration in a maternity care facility: A qualitative study from Botswana | Madisa, Montlenyane et al. | Botswana | 2023 | Qualitative |
| Primary care practice characteristics associated with team functioning in primary care settings in Canada: A practice-based cross-sectional survey | Ndateba, Innocent et al. | Canada | 2023 | Quantitative |
| Perspectives of healthcare professionals toward interprofessional collaboration in primary care settings in a middle eastern country | El-Awaisi, Alla et al. | Qatar | 2021 | Quantitative |
| Midwives' and medical professionals' perspectives of collaborative practice at Queen Elizabeth Central Hospital Maternity unit, Malawi: The discovery phase of an appreciative inquiry project | Chodzaza, Elizabeth et al. | Malawi | 2020 | Qualitative |
| Effect of Relational Characteristics on Management of Wind Farm Interorganizational Construction Projects | Adami, Vivian Sebben et al. | Brazil | 2019 | Qualitative |
| Exploring the importance of team psychological safety in the development of two interprofessional teams | Oleary, Denise Fiona et al. | Ireland | 2016 | Qualitative |
| Facilitators of an Interprofessional Approach to Care in Medical and Mixed Medical/Surgical ICUs: A Multicenter Qualitative Study | Costa, Deena Kelly et al. | United States | 2014 | Qualitative |

Both – Intra/Inter-organizational
| Title | First Author | Country | Publication Year | Study type |
| --- | --- | --- | --- | --- |
| Networks of Care Publication – Manuscript draft |  |  |  |  |
| Improving Care Teams' Functioning: Recommendations from Team Science | Fiscella K et al. | India | 2017 | Analysis |
| Communicating, coordinating, and cooperating when lives depend on it: tips for teamwork | Salas E et al. | Germany | 2008 | Review |
| Teamwork Essentials for Hospitalists | O'Leary KJ et al. | None | 2020 | Analysis |
| Simulation: translation to improved team performance | Hunt EA et al. | USA | 2007 | Review |
| Factors contributing to interprofessional collaboration in Indonesian health centres: A focus group study | Setiadi, Adji Prayitno et al. | Indonesia | 2017 | Qualitative |
| Shared decision-making and interprofessional collaboration in mental healthcare: A qualitative study exploring perceptions of barriers and facilitators | Chong, Wei Wen et al. | Australia | 2013 | Qualitative |
| What are the current barriers to effective cancer care coordination? A qualitative study | Walsh, Jennifer et al. | Australia | 2010 | Qualitative |
| Primary health care in New Zealand: The impact of organisational factors on teamwork | Pullon, Sue et al. | New Zealand | 2009 | Qualitative |
| Difficulties in collaboration: A critical incident study of interprofessional healthcare teamwork | Kvarnström, Susanne | Sweden | 2008 | Qualitative |

Unspecified
| Title | First Author | Country | Publication Year | Study type |
| --- | --- | --- | --- | --- |
| Team development interventions: Evidence-based approaches for improving teamwork | Lacerenza CN | Botswana | 2018 | None |
| Leaving No One Behind: The Role of Primary Care Provider Networks in Advancing Equitable Universal Health Coverage in the South Dayi and South Tongu Districts in Ghana | Ghana Health Service | Ghana | 2021 | None |
| Area-based teamwork for MNCH | The START Center at the University of Washington |  | 2016 | Review |

Based on the scoping review, we also defined organizational enablers for driving teamwork in the context of relational coordination. Organizational enablers can be subdivided into role clarity and standardized processes, performance management, and coordination. For each of the three organizational enablers, we identified core strategies synthesized from the literature review as listed in Figure 6.

**Fig 6.**
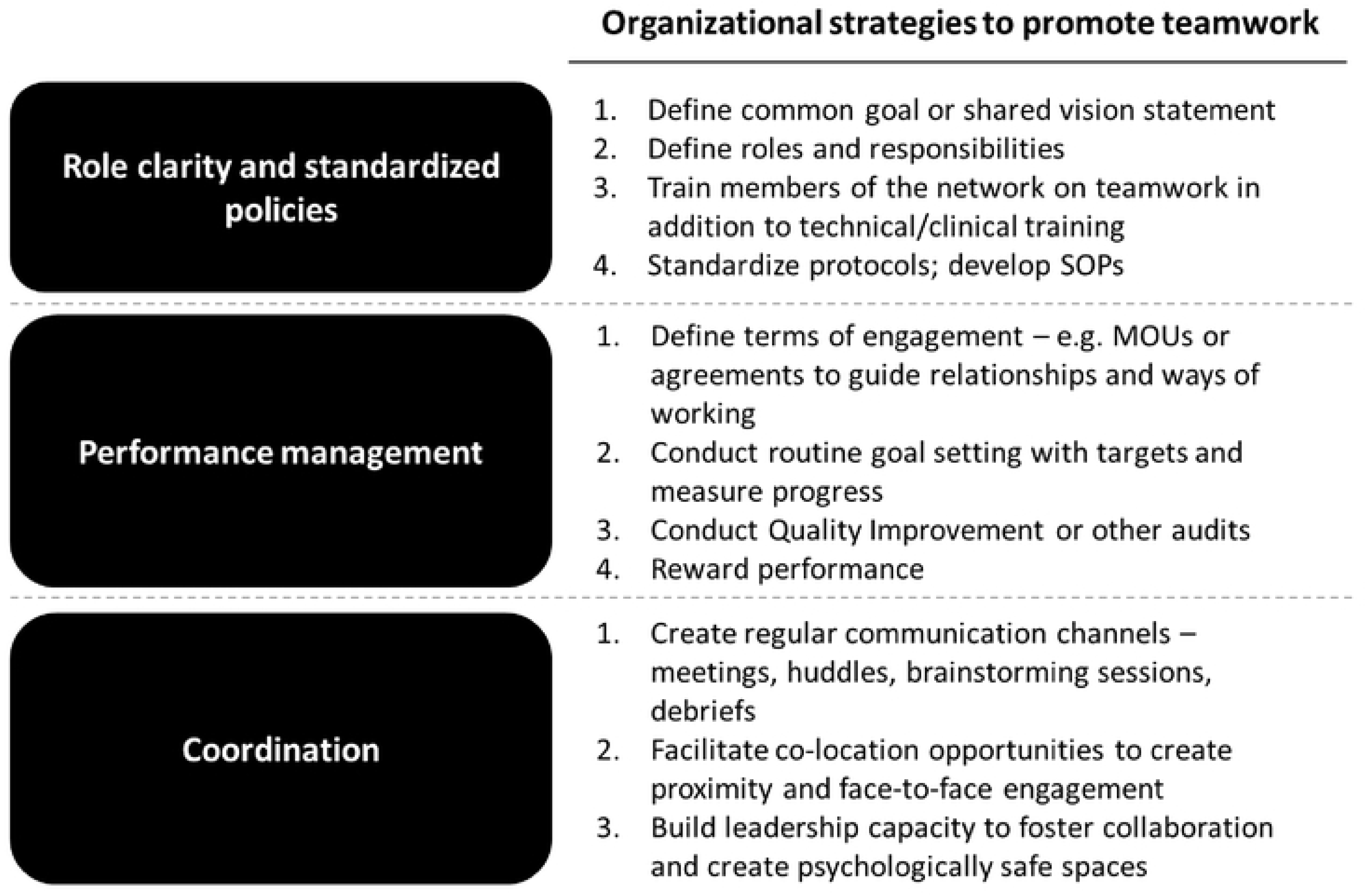
Organizational Enablers to improve teamwork in NOCs

## DISCUSSION

This scoping review examined the relational aspects that define high-performing Networks of Care (NOCs) to develop a revised framework for implementing the NOC model for maternal and newborn health in LMICs. Our broad search strategy, guided by expert opinions, captured a wide range of studies. The cross-disciplinary approach, incorporating business, military, and aviation, underscores the widespread recognition that relational processes drive organizational and team performance. We analyzed 90 relevant papers and found that relational aspects of NOCs for MNH are best described through an adaptation of Relational Coordination theory. The revised framework emphasizes teamwork and organizational enablers for both intra- and inter-organizational networks, built on a strong health system together for successful NOCs. NOCs span clinical and administrative dimensions of care coordination and include teams at the frontlines of healthcare delivery, healthcare managers and administrators, and healthcare service users. Striving for Relational Coordination is not limited to clinical actors but also applies to networks that include administrative and community actors at different levels.

While many attributes and behaviors of teamwork are well-known, some aspects emerged as particularly important for LMICs. First, psychological safety as an attribute may be linked to NOC resilience and adaptive learning. This is supported by existing literature identifying psychological safety as important for improving organizational performance (103) and organizational learning(104), which is crucial for NOCs’ adaptive learning feature. Team psychological safety is a function of interpersonal trust and mutual respect, creating a climate that enables people to speak up and question without fear of judgement (104). Second, trust is built by achieving results, so setting realistic short-term goals rather than overambitious targets is vital. LMICs often align with long-term global targets, but short-term success is critical for NOCs. For LMICs to adopt NOCs, strong and realistic performance management systems are essential. Additionally, collaborative leadership with shared decision-making is necessary to ensure effective service coordination and build trust and transparency.

Careful consideration and implementation of organizational enablers is needed. A dedicated coordinator is key to network success. This role is especially valuable in LMICs, where human resources are limited and professional, cultural, and gender boundaries affect service coordination. Health systems adopting the NOC approach should consider creating boundary-spanning roles as a component of coordination to facilitate operations as they introduce Relational Coordination in NOC.

Finally, a functional healthcare system is needed for effective collaboration and coordination. This includes sufficient infrastructure, appropriate staffing ratios, and adequate funding (21). Efforts to maintain team stability are crucial, as staff attrition and frequent redistribution hinder the development of strong interpersonal relationships necessary for building and maintaining trust which is critical for teamwork sustenance.

A previously implemented system-oriented intervention to improve health outcomes for children in Kenyan district hospitals, the Clinical Information Network (CIN) (105), serves as a useful example to illustrate the applicability of our revised framework. If we apply our framework to describe the CIN intervention package, we can identify components that strengthen teamwork attributes, with alignment around specific shared goals; capacity building included teamwork training specifically to build capacity for collective leadership, and mentorship on both clinical skills and teamwork. Audit and performance review without sanctioning fostered the psychological safety needed for continuous learning and adaptation. Organizational enablers included establishing a functional hospital information system for real-time data on performance with regular feedback mechanisms. The project designating a network coordinator to support the process and facilitate collaboration through meetings and reviews(105); And finally, the project addressed the health system pillars by monitoring and providing equipment, supplies and adequate personnel.

After four years of implementation, the lessons from this CIN intervention showed that resourcing remained a significant threat to the sustainability of the complex and adaptive mechanisms required to keep the network functional and sustained. They also noted that improving care outcomes required more than implementation of a package of interventions, it also relied on building “soft skills” among system stakeholders, leaders, with adaptive learning inbuilt within service delivery processes for success(106).

Our NOC framework overlaps with Mintzberg’s concepts of job specialization, behavior formalization through rules and procedures, and liaison devices for encouraging mutual adjustments across units (107). Healthcare delivery systems are complex, operating across various organizations, disciplines, and contexts. Delivering high-quality, patient-centered services requires reliable collaboration within and across these boundaries (20). Relationships are essential in ensuring integrated, coordinated care in these systems. Collaborative engagements in healthcare range from formal to informal and can be potential, developing, or active (53). Our literature review highlighted more intentional approaches to foster coordination. Often, existing informal relationships are built over time out of necessity and from interpersonal connections (44,53,60,91).

A key takeaway is that Relational Coordination in NOCs in LMICs is highly relevant and with the potential to improve healthcare service delivery if properly understood and applied both within and across institutions, including primarily management and administrative teams designed to support MNH care processes. Clear definitions, measurement approaches, and interventions grounded in a NOC operational framework can help accelerate the introduction of effective interventions to improve MNH services through continuous learning and innovation. In addition, although scarcely explored in LMICs, evidence supports the hypothesis that Relational Coordination is associated with improved job satisfaction, lower burnout, higher motivation, and continuous learning. This can lead to higher retention, addressing a major challenge in healthcare in many LMICs (100,108).

Additional research is needed to develop and test validated tools and measures for evaluating these elements in MNH networks in LMICs. While some existing tools measure teamwork and coordination, they are primarily used in high-income countries and within single organizational units (80,109,110).

Maternal and newborn health is a high-reliability field like aviation, military action, or manufacturing, where processes are complex, and mistakes can be fatal (111,112). An ongoing challenge in healthcare systems, particularly in maternal and newborn health, is the influence of power dynamics and gender on relational coordination processes. Traditional roles and hierarchies within the medical team, comprising doctors, nurses, midwives, and allied health professionals, impact trust and the maintenance of relationships. It is important to go deeper into understanding the roles hierarchy, power, and gender play in the relational elements of NOCs in LMICs through further research.

### Study Strengths and limitations

Our study describes a comprehensive cross-disciplinary approach to develop a revised NOC Operational framework that emphasizes Relational Coordination. Our study did not explore the creation of relational elements or social processes, nor the theory of change for achieving improved performance, although evidence suggests these aspects are important (20,31,113). Our search was limited to primarily published and grey literature available online, acknowledging the possibility of relevant LMIC studies or case studies not publicly accessible which we may have missed. Given the importance of training as a critical organizational enabler for promoting better collaboration and teamwork, the exclusion of articles that studied teamwork training methods may have limited the extent to which we can understand the role of training in NOC models. Finally, while we have extrapolated data from the review and stakeholder consultations to inform the development of the revised operational framework, the paucity of studies and articles from LMICs means that some of the findings may be skewed to high income contexts and additional research on NOCs, and their relational aspects in LMICs may be required to further refine the operational framework.

## CONCLUSION

This study developed a revised operational framework for NOC focusing on Relational Coordination with insights integrated from both healthcare and non-healthcare fields. The resulting NOC framework aims to orient implementers in designing interventions to improve Relational Coordination in maternal and child healthcare services to achieve better health outcomes and build a more resilient workforce. Additional research is needed to develop tools for measuring and evaluating relational coordination in the LMIC context. Of note, power dynamics and gender roles remain significant challenges that are under-explored and less studied in healthcare systems in LMICs, which warrant further inquiry.

## Data Availability

All relevant data are within the manuscript and its Supporting Information files.

## Acknowledgments

The authors would like to thank the stakeholders who were consulted in the process of conducting the literature review and figuring out the strategy,

## Funding acknowledgement

This study was supported by funds from The United States Agency for International Development (USAID) Health Evaluation and Applied Research Development (HEARD) Project.

## Contributors

OF led the scoping review with feedback on the synthesis and analysis from DW, and SB. DW, AJ and OF conducted the stakeholder consultations with inputs from KK, AM, RC, DM, MV. OF drafted the manuscript with support from DW, SB, NP with feedback on subsequent manuscript drafts by all authors.

## Notes

### Competing Interest Statement

The authors have declared no competing interest.

### Funding Statement

Yes

### Author Declarations

This paper did not require IRB approval as no data from human subjects was obtained or analyzed

## References

1. World Health Organization. Improving maternal and newborn health and survival and reducing stillbirth: progress report 2023. [Internet]. Geneva; 2023. Report No.: License: CC BY-NC-SA 3.0 IGO. Available from: https://www.who.int/publications/i/item/9789240073678

2. UN DESA. The Sustainable Development Goals Report 2024. UN DESA New York, USA; 2024 Jun.

3. Kalaris K, Radovich E, Carmone AE, Smith JM, Hyre A, Baye ML, et al. Networks of Care: An Approach to Improving Maternal and Newborn Health. Global Health: Science and Practice [Internet]. 2022 Dec 21 [cited 2023 Oct 19];10(6). Available from: https://www.ghspjournal.org/content/10/6/e2200162

4. Carmone AE, Kalaris K, Leydon N, Sirivansanti N, Smith JM, Storey A, et al. Developing a Common Understanding of Networks of Care through a Scoping Study. Health Syst Reform. 2020 Sep 25;6(2):e1810921.

5. Agyekum EO, Kalaris K, Maliqi B, Moran AC, Ayim A, Roder-DeWan S. Networks of care to strengthen primary healthcare in resource constrained settings. BMJ [Internet]. 2023;380. Available from: https://www.bmj.com/content/380/bmj-2022-071833

6. Gittell JH, Fairfield KM, Bierbaum B, Head W, Jackson R, Kelly M, et al. Impact of relational coordination on quality of care, postoperative pain and functioning, and length of stay: a nine-hospital study of surgical patients. Med Care. 2000 Aug;38(8):807–19.

7. Nickel S, Thiedemann B, von dem Knesebeck O. The effects of integrated inpatient health care on patient satisfaction and health-related quality of life: Results of a survey among heart disease patients in Germany. Health Policy. 2010 Dec;98(2–3):156–63.

8. Sun X, Tang W, Ye T, Zhang Y, Wen B, Zhang L. Integrated care: a comprehensive bibliometric analysis and literature review. Int J Integr Care. 2014 Jun 12;14:e017.

9. Bhatta S, Rajbhandari S, Kalaris K, Carmone AE. The Logarithmic Spiral of Networks of Care for Expectant Families in Rural Nepal: A Descriptive Case Study. Health Syst Reform. 2020 Nov 20;6(2):e1824520.

10. Cordier LF, Kalaris K, Rakotonanahary RJL, Rakotonirina L, Haruna J, Mayfield A, et al. Networks of Care in Rural Madagascar for Achieving Universal Health Coverage in Ifanadiana District. Health Syst Reform. 2020 Sep 1;6(2):e1841437.

11. Fasawe O, Adekeye O, Carmone AE, Dahunsi O, Kalaris K, Storey A, et al. Applying a Client-centered Approach to Maternal and Neonatal Networks of Care: Case Studies from Urban and Rural Nigeria. Health Syst Reform. 2020 Sep 1;6(2):e1841450.

12. Martinez Vergara MT, Angulo de Vera E, Carmone AE. Building Trust to Save Lives in a Metro Manila Public-Private Network of Care: A Descriptive Case Study of Quirino Recognized Partners in Quezon City, Philippines. Health Syst Reform. 2020 Sep 1;6(2):e1815473.

13. Sequeira D’Mello B, Bwile P, Carmone AE, Kalaris K, Magembe G, Masweko M, et al. Averting Maternal Death and Disability in an Urban Network of Care in Dar es Salaam, Tanzania: A Descriptive Case Study. Health Syst Reform. 2020 Sep 1;6(2):e1834303.

14. Saldaña K, Leydon N, Hodgins S. Nurturing and Optimizing Networks of Care to Maximize Benefits to Patients, Health Workers, and Health Systems. Glob Health Sci Pract. 2022 Dec 21;10(6):e2200516.

15. Samuels F, Amaya AB, Balabanova D. Drivers of health system strengthening: learning from implementation of maternal and child health programmes in Mozambique, Nepal and Rwanda. Health Policy and Planning. 2017 Sep 1;32(7):1015–31.

16. Braithwaite J. Bridging gaps to promote networked care between teams and groups in health delivery systems: a systematic review of non-health literature. BMJ Open. 2015 Sep 1;5(9):e006567.

17. Buljac-Samardzic M, Doekhie KD, van Wijngaarden JDH. Interventions to improve team effectiveness within health care: a systematic review of the past decade. Human Resources for Health. 2020 Jan 8;18(1):2.

18. O’Leary KJ, Sehgal NL, Terrell G, Williams MV, High Performance Teams and the Hospital of the Future Project Team. Interdisciplinary teamwork in hospitals: a review and practical recommendations for improvement. J Hosp Med. 2012 Jan;7(1):48–54.

19. Reader TW, Flin R, Mearns K, Cuthbertson BH. Developing a team performance framework for the intensive care unit*. Critical Care Medicine. 2009 May;37(5):1787.

20. Rosen MA, DiazGranados D, Dietz AS, Benishek LE, Thompson D, Pronovost PJ, et al. Teamwork in Healthcare: Key Discoveries Enabling Safer, High-Quality Care. Am Psychol. 2018;73(4):433–50.

21. Wranik WD, Price S, Haydt SM, Edwards J, Hatfield K, Weir J, et al. Implications of interprofessional primary care team characteristics for health services and patient health outcomes: A systematic review with narrative synthesis. Health Policy. 2019 Jun;123(6):550– 63.

22. Grady C, Han H, Kim DH, Coderre-Ball AM, Alam N. Family physicians collaborating for health system integration: a scoping review. BMC Health Services Research. 2023 Jan 23;23(1):68.

23. Karlsson ICM, Mukhtar-Landgren D, Smith G, Koglin T, Kronsell A, Lund E, et al. Development and implementation of Mobility-as-a-Service – A qualitative study of barriers and enabling factors. Transportation Research Part A: Policy & Practice. 2020 Jan;131:283–95.

24. Nicaise P, Grard A, Leys M, Van Audenhove C, Lorant V. Key dimensions of collaboration quality in mental health care service networks. Journal of Interprofessional Care. 2021 Jan 2;35(1):28–36.

25. Gerrish K. Teamwork in primary care: an evaluation of the contribution of integrated nursing teams. Health Soc Care Community. 1999 Sep;7(5):367–75.

26. Madisa M, Filmalter CJ, Heyns T. Healthcare professionals and pregnant and post-natal women’s perceptions of interprofessional collaboration in a maternity care facility: A qualitative study from Botswana. Midwifery. 2023 Oct;125:103768.

27. Portela MC, Lima SML, da Costa Reis LG, Martins M, Aveling EL. Challenges to the improvement of obstetric care in maternity hospitals of a large Brazilian city: an exploratory qualitative approach on contextual issues. BMC Pregnancy and Childbirth. 2018 Nov 26;18(1):459.

28. Spitzer EG, Kaitz J, Fix GM, Harvey KLL, Stadnick NA, Sullivan JL, et al. Developing Relational Coordination: A Qualitative Study of Outpatient Mental Health Teams. Adm Policy Ment Health. 2023 Jul 1;50(4):591–602.

29. Vyt A. Interprofessional and transdisciplinary teamwork in health care. Diabetes Metab Res Rev. 2008;24 Suppl 1:S106–109.

30. Papadimitriou C, Cott C. Client-centred practices and work in inpatient rehabilitation teams: results from four case studies. Disability and Rehabilitation. 2015 Jun 19;37(13):1135–43.

31. Alderwick H, Hutchings A, Briggs A, Mays N. The impacts of collaboration between local health care and non-health care organizations and factors shaping how they work: a systematic review of reviews. BMC Public Health. 2021 Apr 19;21(1):753.

32. Carlini J, Lehman K, Dharmesti M, Knox K. Maximizing value in healthcare partnerships: A case examining an inter-organizational relationship in the public and non-profit sectors. Journal of Philanthropy & Marketing. 2023 Aug;28(3):1–16.

33. Cert KGBMPRRD. Teamwork in primary care: an evaluation of the contribution of integrated nursing teams. Health & Social Care in the Community. 1999;7(5):367–75.

34. de Brito GEG, Benício de Sá JGS, de Andrade AJB, Forte FDS, Batiston AP, Viana da Costa M. Dimensions of interprofessional collaboration in the dynamics of the expanded family health and primary care centers: A qualitative study. J Interprof Care. 2022;36(5):676–83.

35. Santos CM, Uitdewilligen S, Passos AM. Why is Your Team More Creative Than Mine? The Influence of Shared Mental Models on Intra-group Conflict, Team Creativity and Effectiveness. Creativity and Innovation Management. 2015;24(4):645–58.

36. Fiscella K, Mauksch L, Bodenheimer T, Salas E. Improving Care Teams’ Functioning: Recommendations from Team Science. The Joint Commission Journal on Quality and Patient Safety. 2017 Jul 1;43(7):361–8.

37. Leach LS, Myrtle RC, Weaver FA, Dasu S. Assessing the performance of surgical teams. Health Care Management Review. 2009 Mar;34(1):29.

38. Sibbald S, Schouten K, Sedig K, Maskell R, Licskai C. Key characteristics and critical junctures for successful Interprofessional networks in healthcare - a case study. BMC Health Serv Res. 2020 Jul 29;20(1):700.

39. Spigel L, Plough A, Paterson V, West R, Jurczak A, Henrich N, et al. Implementation strategies within a complex environment: A qualitative study of a shared decision-making intervention during childbirth. Birth. 2022;49(3):440–54.

40. Chong WW, Aslani P, Chen TF. Shared decision-making and interprofessional collaboration in mental healthcare: a qualitative study exploring perceptions of barriers and facilitators. J Interprof Care. 2013 Sep;27(5):373–9.

41. Djaharuddin I, Aras I, Masadah R, Yusuf I, Idris I, Rasyid H, et al. Factors Influencing the Implementation of Interprofessional Collaborative Practice in Teaching Hospital Setting: A Mixed-Method Study. J Adv Med Educ Prof. 2023 Oct;11(4):213–21.

42. El Ansari CJP Walid. Partnerships, community participation and intersectoral collaboration in South Africa. Journal of Interprofessional Care. 2001 Jan 1;15(2):119–32.

43. Lacerenza CN, Marlow SL, Tannenbaum SI, Salas E. Team development interventions: Evidence-based approaches for improving teamwork. American Psychologist. 2018;73(4):517– 31.

44. Levesque JF, Harris MF, Scott C, Crabtree B, Miller W, Halma LM, et al. Dimensions and intensity of inter-professional teamwork in primary care: evidence from five international jurisdictions. Fam Pract. 2018 May 23;35(3):285–94.

45. Mitchell C, Tazzyman A, Howard SJ, Hodgson D. More that unites us than divides us? A qualitative study of integration of community health and social care services. BMC Family Practice. 2020 May 29;21(1):96.

46. Peterson WE, Medves JM, Davies BL, Graham ID. Multidisciplinary collaborative maternity care in Canada: easier said than done. J Obstet Gynaecol Can. 2007 Nov;29(11):880–6.

47. Sheppard CL, Gould S, Guilcher SJT, Liu B, Linkewich E, Austen A, et al. “We could be good partners if we work together”: the perspectives of health and social service providers on the barriers to forming collaborative partnerships with social housing providers for older adults. BMC Health Services Research. 2022 Mar 7;22(1):313.

48. Auschra C. Barriers to the Integration of Care in Inter-Organisational Settings: A Literature Review. 2018 Jan 16;18(1):5.

49. El-Awaisi A, Awaisu A, Aboelbaha S, Abedini Z, Johnson J, Al-Abdulla SA. Perspectives of Healthcare Professionals Toward Interprofessional Collaboration in Primary Care Settings in a Middle Eastern Country. J Multidiscip Healthc. 2021;14:363–79.

50. Ervin JN, Kahn JM, Cohen TR, Weingart LR. Teamwork in the Intensive Care Unit. Am Psychol. 2018;73(4):468–77.

51. Palumbo R, Manesh MF, Pellegrini MM, Flamini G. Exploiting Inter-Organizational Relationships in Health Care: A Bibliometric Analysis and Literature Review. Administrative Sciences (2076-3387). 2020 Sep;10(3):57.

52. Christensen LD, Huibers L, Bro F, Christensen MB, Mygind A. Interprofessional team-based collaboration between designated GPs and care home staff: a qualitative study in an urban Danish setting. BMC Primary Care. 2023 Jan 4;24(1):3.

53. Palinkas LA, Fuentes D, Finno M, Garcia AR, Holloway IW, Chamberlain P. Inter-Organizational Collaboration in the Implementation of Evidence-based Practices Among Public Agencies Serving Abused and Neglected Youth. Adm Policy Ment Health. 2014 Jan 1;41(1):74–85.

54. Frykman M, Hasson H, Muntlin Athlin Å, von Thiele Schwarz U. Functions of behavior change interventions when implementing multi-professional teamwork at an emergency department: a comparative case study. BMC Health Services Research. 2014 May 15;14(1):218.

55. Ndateba I, Wong ST, Beaumier J, Burge F, Martin-Misener R, Hogg W, et al. Primary care practice characteristics associated with team functioning in primary care settings in Canada: A practice-based cross-sectional survey. J Interprof Care. 2023;37(3):352–61.

56. Silva IS, Arantes CIS, Fortuna CM. Conflict as a possible catalyst for democratic relations in the work of the Family Health team. Rev esc enferm USP. 2019 Mar 28;53:e03455.

57. van der Lee N, Driessen EW, Scheele F. How the past influences interprofessional collaboration between obstetricians and midwives in the Netherlands: Findings from a secondary analysis. Journal of Interprofessional Care. 2016 Jan 2;30(1):71–6.

58. Downe S, Finlayson K, Fleming A. Creating a collaborative culture in maternity care. J Midwifery Womens Health. 2010;55(3):250–4.

59. Kutash K, Acri M, Pollock M, Armusewicz K, Serene Olin S chin, Hoagwood KE. Quality Indicators for Multidisciplinary Team Functioning in Community-Based Children’s Mental Health Services. Adm Policy Ment Health. 2014 Jan 1;41(1):55–68.

60. Neuhaus C, Lutnæs DE, Bergström J. Emergence of power and complexity in obstetric teamwork. PLOS ONE. 2022 Jun 9;17(6):e0269711.

61. Kerrissey MJ, Singer SJ. Factors affecting collaboration between clinical and community service organizations. Health Care Management Review. 2023 Jun;48(2):130.

62. McCullough JM, Eisen-Cohen E, Lott B. Barriers and facilitators to intraorganizational collaboration in public health: Relational coordination across public health services targeting individuals and populations. Health Care Manage Rev. 2020;45(1):60–72.

63. Aunger JA, Millar R, Greenhalgh J. Modelling lifecycles of inter-organizational collaborations in healthcare: a systematic review and best-fit framework synthesis. J Health Organ Manag. 2023 Aug 2;ahead-of-print(ahead-of-print).

64. Behruzi R, Klam S, Dehertog M, Jimenez V, Hatem M. Understanding factors affecting collaboration between midwives and other health care professionals in a birth center and its affiliated Quebec hospital: a case study. BMC Pregnancy and Childbirth. 2017 Jun 26;17(1):200.

65. Braganza ME. It’s a Relationship: A Qualitative Exploration of the Challenges to Interorganizational Collaborative Relationships. Canadian Journal of Community Mental Health. 2016 Feb 1;35(1):15–28.

66. Zaheer A, McEvily B, Perrone V. Does Trust Matter? Exploring the Effects of Interorganizational and Interpersonal Trust on Performance. Organization Science. 1998 Apr 1;9(2):141–59.

67. Manski-Nankervis JA, Furler J, Blackberry I, Young D, O’Neal D, Patterson E. Roles and relationships between health professionals involved in insulin initiation for people with type 2 diabetes in the general practice setting: a qualitative study drawing on relational coordination theory. BMC Family Practice. 2014 Jan 31;15(1):20.

68. van Dongen JJJ, Lenzen SA, van Bokhoven MA, Daniëls R, van der Weijden T, Beurskens A. Interprofessional collaboration regarding patients’ care plans in primary care: a focus group study into influential factors. BMC Family Practice. 2016 May 28;17(1):58.

69. Williams SP, Purkayastha S, Chaturvedi S, Darzi A. The GP-OH (General Practice – Organizational Health) Survey: Development and Validation of a Novel Instrument to Measure Organizational Health in General Practice. Hospital Topics. 2022 Oct;100(4):177–87.

70. Topp SM, Chipukuma JM. A qualitative study of the role of workplace and interpersonal trust in shaping service quality and responsiveness in Zambian primary health centres. Health Policy Plan. 2016 Mar;31(2):192–204.

71. Degu T, Amsalu E, Kebede A, Adal O. Inter-professional collaboration and associated factors among nurses and physicians in specialized public hospitals, the northwest, Ethiopia: mixed method multi-centered cross-sectional study. BMC Health Services Research. 2023 Mar 27;23(1):286.

72. Leroy L, Rittner JL, Johnson KE, Gerteis J, Miller T. Facilitative Components of Collaborative Learning: A Review of Nine Health Research Networks. Healthc Policy. 2017 Feb;12(3):19–33.

73. Acri MC, Palinkas L, Hoagwood KE, Shen S, Schoonover D, Reutz JR, et al. Interorganizational Relationships among Family Support Organizations and Child Mental Health Agencies. Adm Policy Ment Health. 2014 Jul;41(4):447–54.

74. Setiadi AP, Wibowo Y, Herawati F, Irawati S, Setiawan E, Presley B, et al. Factors contributing to interprofessional collaboration in Indonesian health centres: A focus group study. Journal of Interprofessional Education & Practice. 2017 Sep 1;8:69–74.

75. Szafran O, Torti JMI, Kennett SL, Bell NR. Family physicians’ perspectives on interprofessional teamwork: Findings from a qualitative study. Journal of Interprofessional Care. 2018 Mar 4;32(2):169–77.

76. Wise S, Duffield C, Fry M, Roche M. A team mental model approach to understanding team effectiveness in an emergency department: A qualitative study. J Health Serv Res Policy. 2022 Jan;27(1):14–21.

77. Bolton R, Logan C, Gittell JH. Revisiting Relational Coordination: A Systematic Review. The Journal of Applied Behavioral Science. 2021 Sep 1;57(3):290–322.

78. Dynes MM, Stephenson R, Hadley C, Sibley LM. Factors shaping interactions among community health workers in rural Ethiopia: rethinking workplace trust and teamwork. J Midwifery Womens Health. 2014 Jan;59 Suppl 1:S32–43.

79. Clark W. Teamwork: A Multidisciplinary Review. Child Adolesc Psychiatr Clin N Am. 2021 Oct;30(4):685–95.

80. Jaruseviciene L, Liseckiene I, Valius L, Kontrimiene A, Jarusevicius G, Lapão LV. Teamwork in primary care: perspectives of general practitioners and community nurses in Lithuania. BMC Family Practice. 2013 Aug 15;14(1):118.

81. Karlsson M, Garvare R, Zingmark K, Nordström B. Organizing for sustainable inter-organizational collaboration in health care processes. Journal of Interprofessional Care. 2020 Mar 3;34(2):241–50.

82. Howell EA, Sofaer S, Balbierz A, Kheyfets A, Glazer KB, Zeitlin J. Distinguishing High-Performing from Low-Performing Hospitals for Severe Maternal Morbidity: A Focus on Quality and Equity. Obstet Gynecol. 2022 Jun 1;139(6):1061–9.

83. Mahmoud Z, Angelé-Halgand N, Churruca K, Ellis LA, Braithwaite J. The impact of lean management on frontline healthcare professionals: a scoping review of the literature. BMC Health Services Research. 2021 Apr 26;21(1):383.

84. Radnor ZJ, Holweg M, Waring J. Lean in healthcare: The unfilled promise? Social Science & Medicine. 2012 Feb 1;74(3):364–71.

85. Merry AF, Weller J, Mitchell SJ. Teamwork, Communication, Formula-One Racing and the Outcomes of Cardiac Surgery. J Extra Corpor Technol. 2014 Mar;46(1):7–14.

86. Andersen PO, Jensen MK, Lippert A, Østergaard D. Identifying non-technical skills and barriers for improvement of teamwork in cardiac arrest teams. Resuscitation. 2010 Jun;81(6):695–702.

87. Hunt EA, Shilkofski NA, Stavroudis TA, Nelson KL. Simulation: translation to improved team performance. Anesthesiol Clin. 2007 Jun;25(2):301–19.

88. Bentley M, Freeman T, Baum F, Javanparast S. Interprofessional teamwork in comprehensive primary healthcare services: Findings from a mixed methods study. J Interprof Care. 2018 May;32(3):274–83.

89. Doekhie KD, Buljac-Samardzic M, Strating MMH, Paauwe J. Who is on the primary care team? Professionals’ perceptions of the conceptualization of teams and the underlying factors: a mixed-methods study. BMC Fam Pract. 2017 Dec 28;18(1):111.

90. Costa DK, Barg FK, Asch DA, Kahn JM. Facilitators of an interprofessional approach to care in medical and mixed medical/surgical ICUs: a multicenter qualitative study. Res Nurs Health. 2014 Aug;37(4):326–35.

91. Surendran S, Foo CD, Matchar DB, Ansah JP, Car J, Koh GCH. Developing integration among stakeholders in the primary care networks of Singapore: a qualitative study. BMC Health Services Research. 2022 Jun 15;22(1):782.

92. Van Houdt S, Sermeus W, Vanhaecht K, De Lepeleire J. Focus groups to explore healthcare professionals’ experiences of care coordination: towards a theoretical framework for the study of care coordination. BMC Fam Pract. 2014 Dec 24;15:177.

93. McInnes E, Middleton S, Gardner G, Haines M, Haertsch M, Paul CL, et al. A qualitative study of stakeholder views of the conditions for and outcomes of successful clinical networks. BMC Health Serv Res. 2012 Feb 28;12:49.

94. Epps HR, Levin PE. The TeamSTEPPS Approach to Safety and Quality. Journal of Pediatric Orthopaedics [Internet]. 2015;35. Available from: https://journals.lww.com/pedorthopaedics/fulltext/2015/07001/the_teamstepps_approach_to_safety_and_quality.11.aspx

95. Weaver SJ, Rosen MA, DiazGranados D, Lazzara EH, Lyons R, Salas E, et al. Does teamwork improve performance in the operating room? A multilevel evaluation. Jt Comm J Qual Patient Saf. 2010 Mar;36(3):133–42.

96. Burns LR, Nembhard IM, Shortell SM. Integrating network theory into the study of integrated healthcare. Social Science & Medicine. 2022 Mar 1;296:114664.

97. Bravi F, Gibertoni D, Marcon A, Sicotte C, Minvielle E, Rucci P, et al. Hospital network performance: A survey of hospital stakeholders’ perspectives. Health Policy. 2013 Feb;109(2):150–7.

98. Gittell JH. Relational Coordination. In: Wiley Encyclopedia of Management [Internet]. John Wiley & Sons, Ltd; 2015 [cited 2024 Dec 5]. p. 1–6. Available from: https://onlinelibrary.wiley.com/doi/abs/10.1002/9781118785317.weom110025

99. Falatah R, Conway E. Linking relational coordination to nurses’ job satisfaction, affective commitment and turnover intention in Saudi Arabia. J Nurs Manag. 2019 May;27(4):715–21.

100. House S, Wilmoth M, Kitzmiller R. Relational coordination and staff outcomes among healthcare professionals: a scoping review. J Interprof Care. 2022;36(6):891–9.

101. Hustoft M, Biringer E, Gjesdal S, Aβmus J, Hetlevik Ø. Relational coordination in interprofessional teams and its effect on patient-reported benefit and continuity of care: a prospective cohort study from rehabilitation centres in Western Norway. BMC Health Serv Res. 2018 Sep 17;18(1):719.

102. Havens DS, Vasey J, Git℡l JH, Lin WT. Relational coordination among nurses and other providers: impact on the quality of patient care. Journal of Nursing Management. 2010;18(8):926–37.

103. Schulte M, Cohen NA, Klein KJ. The Coevolution of Network Ties and Perceptions of Team Psychological Safety. Organization Science. 2012 Apr;23(2):564–81.

104. Edmondson A. Psychological Safety and Learning Behavior in Work Teams. Administrative Science Quarterly. 1999 Jun 1;44(2):350–83.

105. English M. Designing a theory-informed, contextually appropriate intervention strategy to improve delivery of paediatric services in Kenyan hospitals. Implement Sci. 2013 Mar 28;8:39.

106. Irimu G, Ogero M, Mbevi G, Agweyu A, Akech S, Julius T, et al. Approaching quality improvement at scale: a learning health system approach in Kenya. Arch Dis Child. 2018 Nov;103(11):1013–9.

107. Mintzberg H. Structure in 5’s: A Synthesis of the Research on Organization Design. Management Science. 1980;26(3):322–41.

108. Van Osch M, Scarborough K, Crowe S, Wolff AC, Reimer-Kirkham S. Understanding the factors which promote registered nurses’ intent to stay in emergency and critical care areas. J Clin Nurs. 2018 Mar;27(5–6):1209–15.

109. Attell BK, Kingery K, Adimu T, Butts J, Howard P, Saha S, et al. Development and Validation of a Measure to Assess Readiness to Advance Health and Equity: The Assessment for Advancing Community Transformation (AACT). Eval Health Prof. 2023 Dec;46(4):309–19.

110. de Wet C, Johnson P, Mash R, McConnachie A, Bowie P. Measuring perceptions of safety climate in primary care: a cross-sectional study. J Eval Clin Pract. 2012 Feb;18(1):135–42.

111. Gaba DM. Structural and Organizational Issues in Patient Safety: A Comparison of Health Care to other High-Hazard Industries. California Management Review. 2000 Oct 1;43(1):83– 102.

112. Leape LL. Error in Medicine. JAMA. 1994 Dec 21;272(23):1851–7.

113. Will K, Johnson M, Lamb G. Team-Based Care and Patient Satisfaction in the Hospital Setting: A Systematic Review. Journal of Patient-Centered Research and Reviews. 2019 Apr 29;6(2):158–71.

